# Use of systemic menopausal hormone therapy among French women: a descriptive nationwide study over the 2011 to 2024 period

**DOI:** 10.64898/2026.09.07.26362441

**Authors:** Ana-Maria Vilcu, Perrine Goussault-Capmas, Yue Zhai, Marianne Canonico, Arnaud Fauconnier, Jacques Bénichou, Lucas Morin, Vivian Viallon, Agnès Fournier, Anne C.M. Thiébaut

## Abstract

**Background:** Worldwide use of systemic menopausal hormone therapy (MHT) declined sharply after 2002 following reports of increased breast cancer and cardiovascular risks. Evidence that the benefit-risk balance depends on initiation timing and treatment patterns has renewed clinical interest in MHT recently. A comprehensive characterisation of utilisation patterns is thus needed to assess alignment with guidelines. We describe systemic MHT use among women aged ≥45 years in France between 2011 and 2024.

**Methods:** In this descriptive nationwide study using the French National Health Data System (SNDS), we included 21,983,503 women aged ≥45 years affiliated with the main health insurance scheme between 2011 and 2024. Systemic MHT was identified from reimbursements for drugs containing systemic oestrogen. Users had at least two reimbursements for systemic oestrogen (unopposed or with a progestogen) within 12 months, and initiators had no reimbursements in the preceding five years. Annual prevalence, initiation and discontinuation proportions were estimated overall and by age group.

**Results:** We identified 1,250,037 systemic MHT users and 578,668 initiators between 2011 and 2024. Annual prevalence decreased from 4.3% in 2011 to 2.0% in 2022, then stabilised, with a slight increase after 2022 among women aged 45-59 years. Prevalence was consistently highest among women aged 55-59 years. Initiation declined from 0.5% of women in 2011 to 0.2% in 2022, before increasing to 0.3% in 2024, and was highest among women aged 50-54 years. Discontinuation was highest among women aged 45-49 years, peaking in 2019, and remained stable in older age groups. Median age at initiation was 52 (Interquartile Range [IQR] 50-55) years, and median treatment duration was 2.1 (IQR 0.6-6.1) years. Most initiators (79.3%) started combined oestrogen-progestogen therapy. Oestradiol was used almost exclusively (98.5%), mainly transdermally (74.2%). Micronised progesterone was the predominant progestogen, used exclusively by 62.9% of combined therapy initiators, followed by dydrogesterone (9.2%).

**Conclusion:** After prolonged declining use, systemic MHT initiation increased modestly after 2022. Prescribing appeared broadly aligned with current recommendations favouring transdermal oestradiol combined with micronised progesterone, for relatively short durations. These findings provide a benchmark for public health authorities to monitor alignment between prescribing patterns and guidelines and inform future decisions.

## Background

At menopause, the definitive decline in ovarian oestrogen and progesterone production may trigger climacteric symptoms, including vasomotor symptoms (e.g., hot flushes and night sweats) and the genitourinary syndrome of menopause (e.g., vaginal dryness and urinary troubles).[1, 2] Management of these symptoms includes vaginal oestrogens for genitourinary syndrome of menopause, or systemic menopausal hormone therapy (MHT), which effectively alleviates both by restoring systemic oestrogen levels.[1–5] Systemic MHT consists of oestrogen, administered orally or transdermally (patches or gels), combined with a progestogen in women with an intact uterus to reduce the risk of endometrial cancer associated with unopposed oestrogen.[6]

The publication of the US Women’s Health Initiative (WHI) randomised controlled trial in 2002 and the UK Million Women Study cohort in 2003 fundamentally reshaped systemic MHT prescribing guidelines worldwide and led to a sharp decline in MHT use after 2003, observed at least until the early 2010s.[7–12] The WHI trial reported an unfavourable benefit-risk balance for the specific regimen evaluated in the trial (oral conjugated equine oestrogens combined with medroxyprogesterone acetate). The Million Women Study confirmed an increased breast cancer risk with other regimens as well.[7–10]

Subsequent studies have shown that MHT-associated risks vary according to the treatment regimen, route of oestrogen administration, timing of initiation, duration of use, and type of progestogen, and may attenuate after treatment discontinuation.[11, 13] These findings have prompted a reassessment of the WHI conclusions, particularly regarding the specific regimen investigated and the age of the trial participants. This evolving evidence has renewed clinical interest in MHT, making a comprehensive characterisation of contemporary utilisation patterns both timely and relevant, e.g., for evaluating adherence to current prescribing recommendations.

Yet, contemporary utilisation patterns remain poorly characterised in most countries. While some recent studies reported heterogeneous trends over the past decade, from continued decline in the US and South Korea to recent increases in the UK and Sweden, treatment patterns were rarely described.[14–18] In France, where transdermal oestrogens are preferred over oral formulations and micronised progesterone as the progestogen component of combined MHT has long represented a distinct feature of MHT prescribing,[19–21] recent reports have documented a continuous decline in systemic MHT use between 2001 and 2022,[23, 24] followed by a recent increase.[24] However, these reports provide limited information on treatment duration, treatment switching and discontinuation, prescribing patterns, and the characteristics of women who initiated MHT before 2024.

We therefore aimed to provide a comprehensive description of systemic MHT use in France between 2011 and 2024. The primary objective was to characterise utilisation patterns among women aged 45 years or older initiating systemic MHT, including age at initiation, treatment duration, regimens, progestogen type, routes of oestrogen administration, and treatment changes over time. The secondary objective was to estimate annual proportions of prevalent users, initiators, and discontinuers and to describe temporal trends in these indicators.

## Methods

### Data source

This study used the French National Health Data System (SNDS, *Système National des Données de Santé)* which covers 98.8% of the French population (>66 million individuals), and contains anonymised individual-level information on: (i) sociodemographic characteristics, (ii) outpatient health care reimbursements, (iii) hospital discharge summaries, and iv) long-term disease registrations entitling to full reimbursement of health care.[22] Medical diagnoses are coded using the International Classification of Diseases, 10th Edition (ICD-10). Dispensed drugs are identified through a France-specific pharmaceutical product code (CIP – *Code Identifiant de Présentation*), linked to the corresponding Anatomical Therapeutic Chemical (ATC) code. Drug indications and prescribed daily dosages are not available.

### Study population and design

Women in the SNDS enrolled in the main insurance scheme were eligible for inclusion from January 1^st^ of their 45^th^ anniversary year or January 1^st^, 2011, whichever occurred last, until December 31^st^, 2024, or their date of death, whichever occurred first.

Among women eligible for inclusion* (terms followed by * are defined in Table 1), we performed: (i) annually repeated cross-sectional analyses to estimate annual MHT prevalence*, initiation*, and discontinuation* proportions between 2011 and 2024, and (ii) a longitudinal cohort study focused on systemic MHT initiators* identified between 2011 and 2024, followed from treatment initiation (index date*) until December 31^st^, 2025 or death, whichever occurred first, to describe MHT utilisation patterns.

**Table 1.**
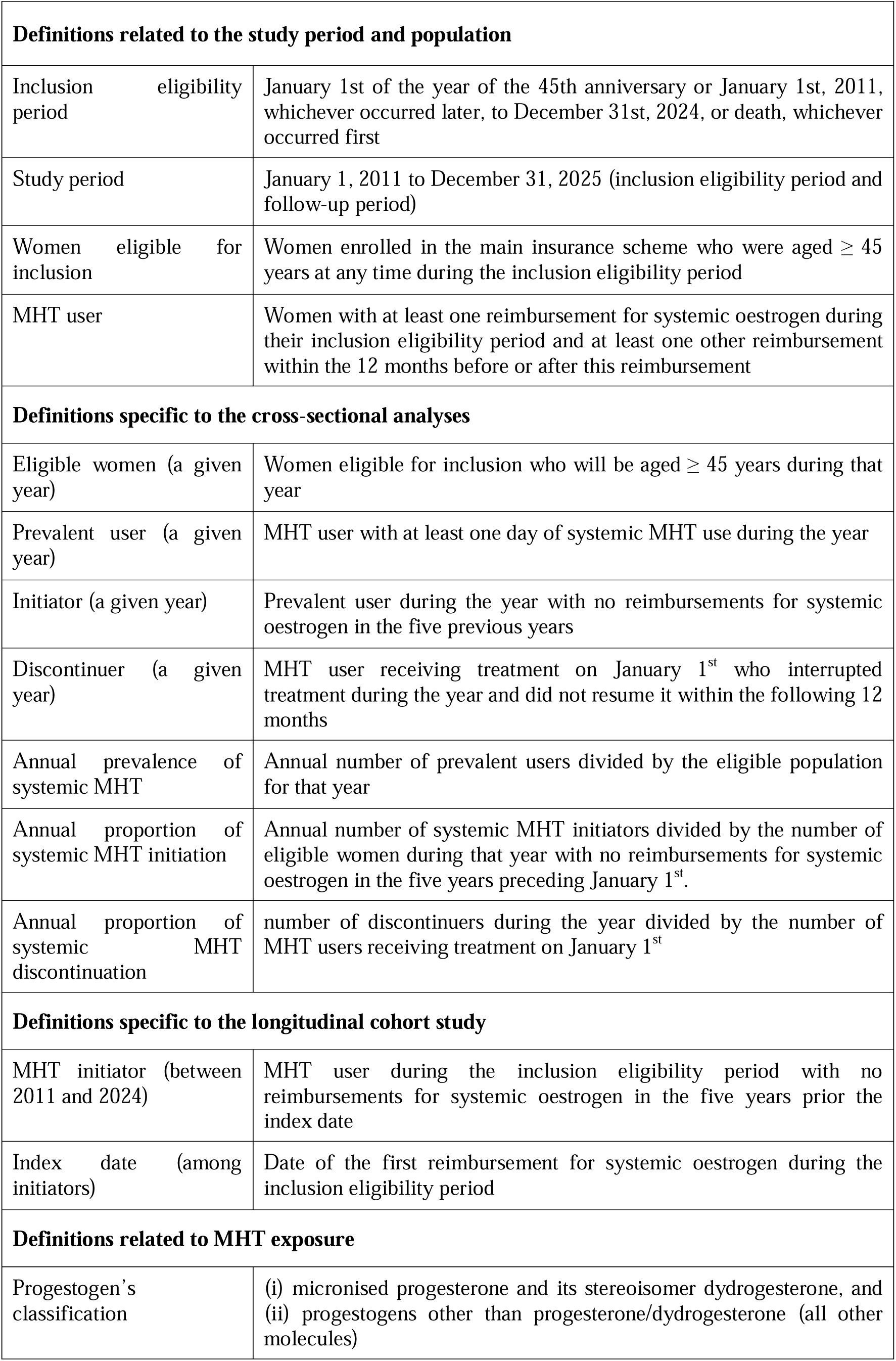
Definitions.

This study is reported in accordance with the RECORD-PE guidelines for cross-sectional and cohort studies using observational routinely collected health data for pharmacoepidemiology.[23]

### Assessment of systemic MHT use

Systemic MHT use was assessed using reimbursements for drugs containing systemic oestrogen from the ATC classes G03C (oestrogens), G03F (progestogens and oestrogens in combination), and G03HB01 (cyproterone and oestrogen). To be considered MHT users*\**, women were required to have at least one reimbursement for systemic oestrogen during their inclusion eligibility period* and at least one other reimbursement within the 12 months before or after this reimbursement. This criterion was used to exclude isolated reimbursements that were unlikely to reflect actual MHT use. We then identified continuous treatment episodes, assuming a 30-day supply per box for oral pills or transdermal patches (based on drug leaflet information) and a 60-day supply per tube of transdermal gels. We assumed no leftovers (full intake between refills) and allowed a 90-day grace period (maximum treatment discontinuation between successive refills) to account for potential dosage adjustments or refill irregularities.

### Assessment of MHT regimen

In France, MHT combining oestrogen and progestogen may consist of fixed-dose combination products containing both hormones (ATC classes G03F and G03HB01) or of separate pharmaceutical products for each hormone. Thus, to distinguish unopposed oestrogen therapy from combined therapy, we additionally searched for reimbursements for progestogens in ATC class G03D (excluding ATC code G03DB08 – dienogest, not indicated for MHT) occurring within three months before or after each oestrogen reimbursement. Based on their structural proximity to natural progesterone, progesterone receptor selectivity, and their potentially distinct risk profiles for breast,[24] endometrial,[25] and ovarian cancers,[26] as well as cardiovascular diseases,[27–30] progestogens were further classified* into micronised progesterone and its stereoisomer dydrogesterone, and other synthetic progestogens.

### Assessment of individual characteristics

Medical events were identified using ICD-10 codes for hospital diagnoses or long-term disease registrations, medical procedure codes, ATC codes, or diagnosis-related groups, validated with gynaecologists or from algorithms previously developed for this database.[31–36]

We assessed, during the five years preceding the index date*, a history of conditions representing potential contraindications or barriers to systemic MHT prescribing: gynaecological cancers (breast, ovarian, uterine), other cancers, venous thromboembolism (VTE), and cardio-neurovascular diseases other than VTE.[31, 32] Outpatient healthcare utilisation at the index date* was measured as the number of days with at least one visit in general practice, gynaecology, or another medical speciality in the prior 12 months, while recent screening mammography (**Additional File 1 eTable 1**) was evaluated within the prior 6 months.

During the five years prior the index date* and throughout follow-up, we assed hysterectomy (which influences the prescribed MHT regimen), bilateral oophorectomy (which induces menopause), infertility treatments and severe endometriosis (alternative indications for systemic oestrogens) (event identification codes shown in **Additional File 1 eTables 1**). We further identified reimbursements for: (i) osteoporosis treatments **(Additional File 1 eTable 1)**, (ii) gonadotropin-releasing hormone agonists (GnRHa; ATC classes L02AE04 and L02AE02), (iii) levonorgestrel-releasing intrauterine system (LNG-IUS; ATC class G02BA03), and (iv) vaginal oestrogens (products administered vaginally from ATC classes G03C, G03F, and G01AC03). GnRHa, indicated for benign conditions such as endometriosis and uterine fibroids, inhibit oestrogen production and may induce menopausal-like symptoms requiring hormonal add-back therapy.[37] LNG-IUS maintained after MHT initiation may provide local progestogenic protection,[38] while vaginal oestrogens may be prescribed to treat the genitourinary syndrome of menopause.

### Statistical analysis

As this was a descriptive study, no prespecified hypothesis was formulated, and no formal null-hypothesis testing was performed.

The annual prevalence*, initiation proportion*, and discontinuation proportion* of systemic MHT were calculated overall among women aged 45 years or older, by 5-year age groups, and by oestrogen administration route and MHT regimen.

We described the individual characteristics of MHT initiators* at the index date*, and compared them with those of a reference population of women not initiating systemic MHT. The reference population was constructed using 1:4 risk-set sampling without replacement. Specifically, for each initiator, up to four women born in the same calendar year who had no reimbursement for systemic oestrogens on the index date or during the preceding five years were randomly selected and assigned the same index date as their matched initiator. This approach ensured comparable timing for the assessment of characteristics in initiators and non-initiators.

Treatment-related characteristics of MHT initiators aged 45 years or older, including regimen, hormone type, and oestrogen administration routes throughout follow-up, were described. Because women could change treatment during follow-up, we estimated the proportion of person-months spent under each treatment regimen, oestrogen administration route, and progestogen class. The cumulative duration of treatment episodes, the duration of the first treatment episode, the time between treatment initiation and the last observed use, and the number of treatment episodes during follow-up were analysed using the Kaplan-Meier method. Treatment discontinuation was considered the event of interest, and women who were still receiving systemic MHT at the end of follow-up were censored. The median (and interquartile range - IQR) pause between the first two exposure episodes was assessed among MHT initiators with multiple episodes.

We conducted a subgroup analysis restricted to women aged 50-59 years, among which systemic MHT use is most prescribed.

All statistical analyses were performed using R (version 4.3.3) and SAS Enterprise Guide (version 8).

## Results

Between 2011 and 2024, we identified 1,250,037 systemic MHT users among 21,983,503 eligible women aged 45 years or older. A total of 578,668 women initiated treatment during this period and were included in the longitudinal cohort study focusing on MHT initiators (**Figure 1**).

**Figure 1:**
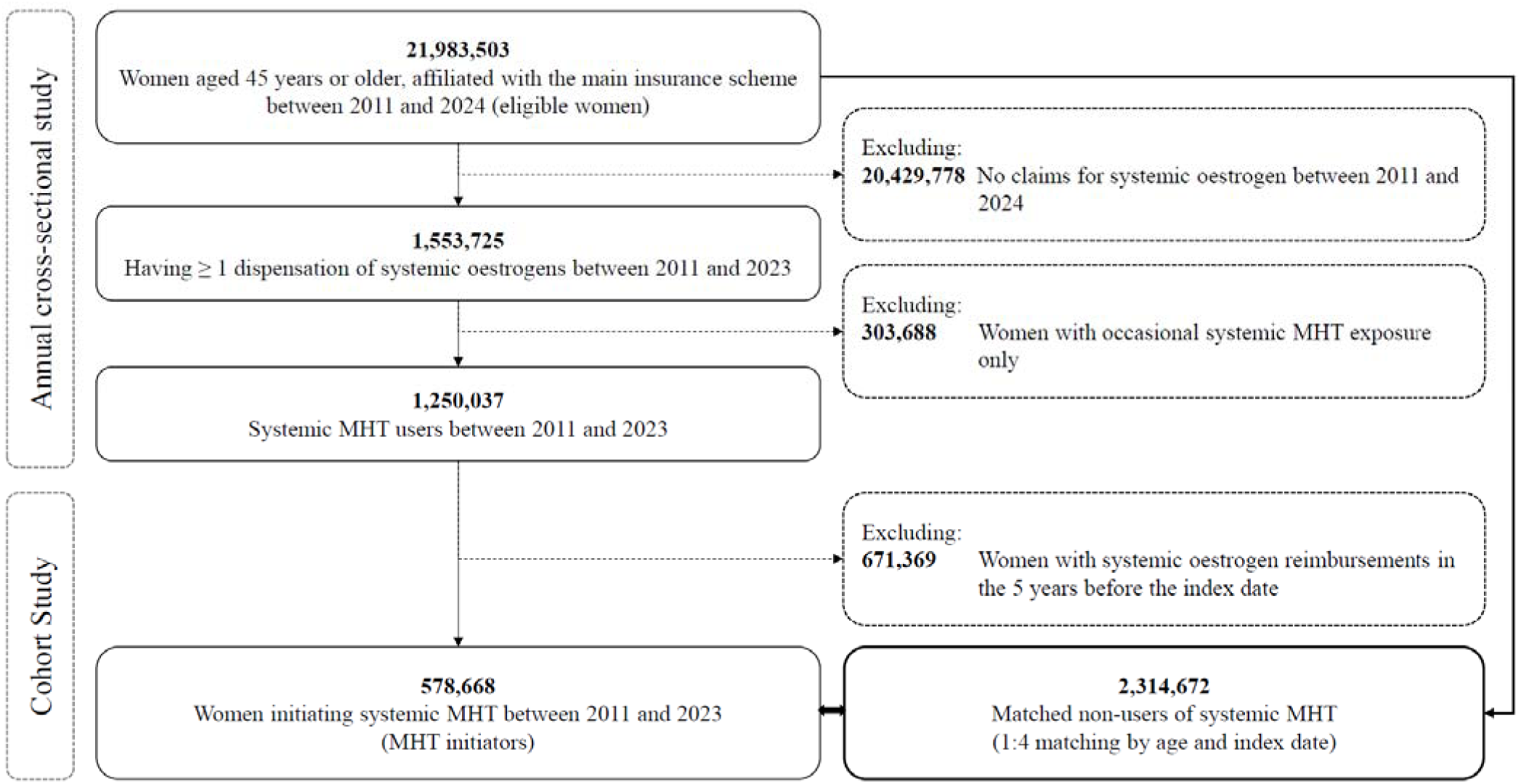
Flowchart of identification of MHT users

### Annual prevalence, initiation and discontinuation of systemic MHT

Among women aged 45 years or older, the annual prevalence of systemic MHT decreased from 4.3% in 2011 to 2.0% in 2022, and remained stable thereafter (**Table 2)**. Similar trends were observed for all routes of oestrogen administration and MHT regimens. The lowest prevalence throughout the period and the largest relative decreases were observed for MHT containing oral oestrogens (-72%), and among regimens combining oestrogen with progestogens other than progesterone or dydrogesterone (-91%) **(Additional File 1 eFigure 1)**. MHT levels and trends varied by age. Prevalence was highest among women aged 55-59 years, followed by women aged 50-54 years and 60-64 years. After 2022, it increased slightly among women aged 45-59 years, and continued to decrease among older age groups (**Figure 2A)**.

**Figure 2.**
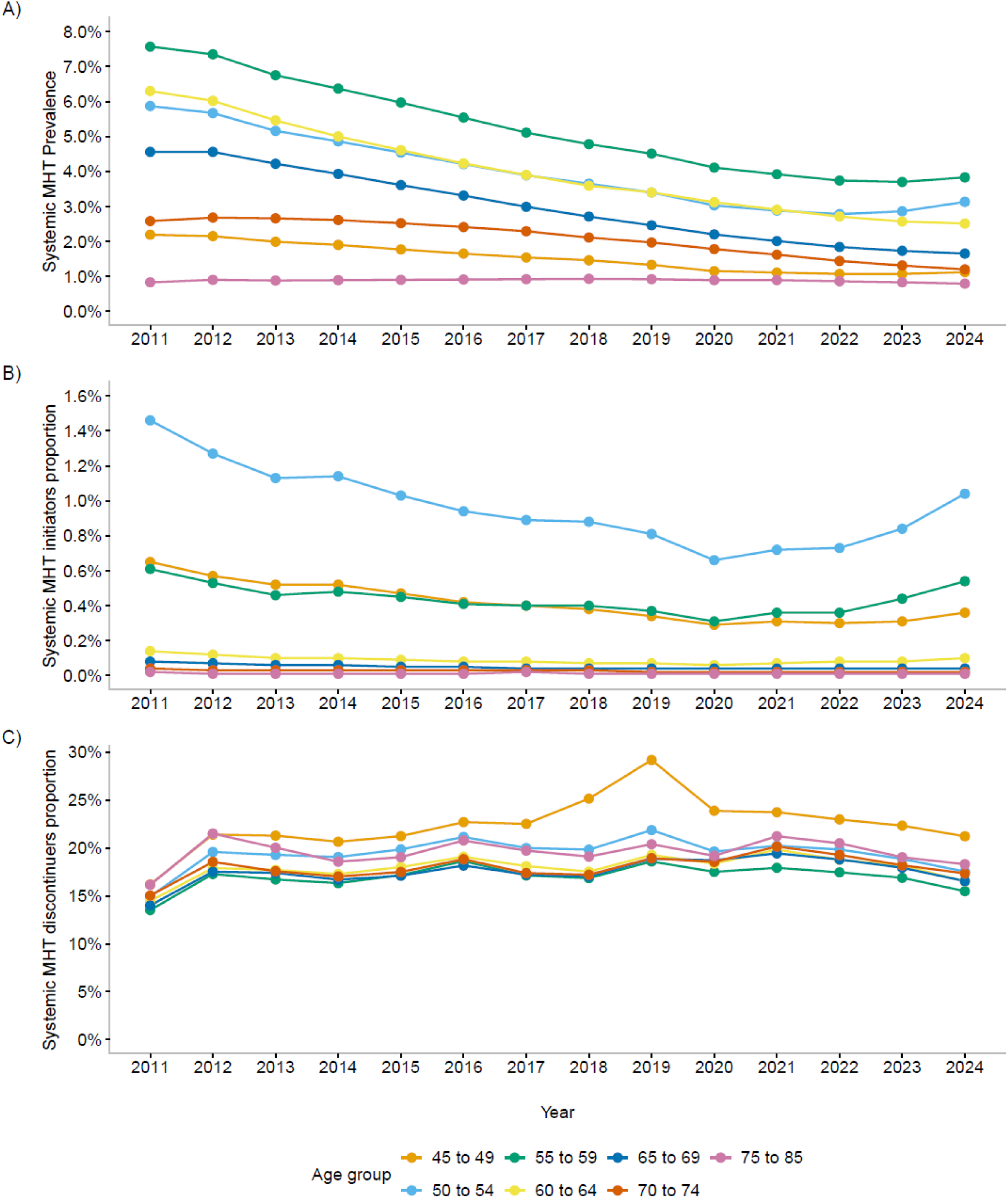
Annual prevalence, initiation, and discontinuation proportions of systemic MHT between 2011 and 2024. **Panel A)** - Annual prevalence of systemic MHT; **Panel B)** – Annual proportion of MHT initiators; **Panel C)** Annual proportion of MHT discontinuers

**Table 2.** Annual prevalence, initiation, and discontinuation proportions of systemic MHT, from 2011 to 2024.

| Year | Prevalence of systemic MHT |  |  | Proportion of systemic MHT initiation |  |  | Proportion of systemic MHT discontinuation |  |  |
| --- | --- | --- | --- | --- | --- | --- | --- | --- | --- |
|  | Systemic MHT prevalent users | Total eligible women | % | Systemic MHT initiators | Population at risk* | % | Systemic MHT discontinuers | Systemic MHT users on January 1st | % |
| 2011 | 615,078 | 14,452,528 | 4.3 | 59,290 | 12,795,520 | 0.5 | 62,461 | 432,898 | 14.4 |
| 2012 | 608,641 | 14,621,012 | 4.2 | 52,060 | 13,054,467 | 0.4 | 83,577 | 454,402 | 18.4 |
| 2013 | 568,281 | 14,789,818 | 3.8 | 47,220 | 13,324,609 | 0.4 | 74,096 | 411,887 | 18.0 |
| 2014 | 541,604 | 14,957,507 | 3.6 | 48,132 | 13,585,762 | 0.4 | 68,964 | 394,199 | 17.5 |
| 2015 | 512,473 | 15,131,001 | 3.4 | 44,118 | 13,812,918 | 0.3 | 68,249 | 375,515 | 18.2 |
| 2016 | 481,365 | 15,297,334 | 3.2 | 40,702 | 14,040,807 | 0.3 | 68,845 | 353,571 | 19.5 |
| 2017 | 450,252 | 15,471,270 | 2.9 | 39,230 | 14,268,460 | 0.3 | 57,229 | 311,814 | 18.4 |
| 2018 | 423,646 | 15,634,915 | 2.7 | 38,387 | 14,492,766 | 0.3 | 53,006 | 290,707 | 18.2 |
| 2019 | 399,389 | 15,788,263 | 2.5 | 35,752 | 14,687,801 | 0.2 | 57,929 | 287,847 | 20.1 |
| 2020 | 364,404 | 15,921,340 | 2.3 | 29,696 | 14,870,926 | 0.2 | 49,008 | 260,637 | 18.8 |
| 2021 | 345,065 | 16,045,115 | 2.2 | 32,673 | 15,043,813 | 0.2 | 48,206 | 244,759 | 19.7 |
| 2022 | 327,234 | 16,180,295 | 2.0 | 33,454 | 15,224,349 | 0.2 | 43,183 | 226,969 | 19.0 |
| 2023 | 320,717 | 16,317,512 | 2.0 | 38,071 | 15,402,296 | 0.3 | 39,882 | 218,727 | 18.2 |
| 2024 | 325,857 | 16,482,884 | 2.0 | 46,316 | 15,594,710 | 0.3 | 36,363 | 214,757 | 16.9 |

\* Population at risk = potential initiators, i.e., non-initiators on January 1<sup>st</sup> with no reimbursements for systemic oestrogens in the five previous years

The annual proportion of systemic MHT initiators aged 45 years or older declined steadily from 0.5% in 2011 to 0.2% in 2022, before increasing to 0.3% in 2024 **(Table 2)**. Increases were limited to MHT containing transdermal (but not oral) oestrogens and to regimens combining oestrogen with micronised progesterone or dydrogesterone **(Additional File 1 eFigure 2)**. MHT initiation was highest among women aged 50-54 years, followed by those aged 55-59 and 45-49 years. After 2022, it increased among women younger than 65 years and remained low and stable in older age groups **(Figure 2B)**.

The annual proportion of discontinuers among users aged 45 years or older was 14.4% in 2011, fluctuated between 17.5% and 20.1% between 2012 and 2023, and decreased to 16.9% in 2024 **(Table 2)**. Discontinuation was slightly higher among women aged 45-49 years, with a peak observed in 2019, notably for orally administered and unopposed oestrogens **(Figure 2C and Additional File 1 eFigures 3-4)**.

Among women aged 50-59 years specifically, the prevalence decreased from 6.7% in 2011 to 3.2% in 2022, before increasing slightly to 3.5% in 2024. The proportion of MHT initiators decreased from 1.1% in 2011 to 0.5% in 2021, increasing to 0.8% in 2024 **(Additional File 1 eFigures 6-8)**. Discontinuation patterns were similar to those observed among women aged 45 years or older.

### Characteristics of MHT initiators aged 45 years or older at the index date

Among the 578,668 systemic MHT initiators, 20.7% (n=117,303) started with unopposed oestrogens and 79.3% (n=461,365) with combined oestrogen-progestogen therapy (**Table 3**). The median age at initiation was 52 [IQR 50-55] years and was similar across the two regimens. Compared with the reference population of non-initiators, initiators of both unopposed and combined MHT had higher healthcare utilisation in the 12 months before the index date, including more frequent visits to general practitioners, gynaecologists, and other healthcare professionals. They were also more likely to have undergone recent mammography and to have a history of vaginal oestrogen or GnRHa use, and less likely to have a history of breast cancer or other cardio-neurovascular diseases **(Table 3)**. Compared with both the reference population of MHT non-initiators and initiators of combined MHT, women initiating unopposed oestrogen therapy were more likely to have had a hospitalisation for hysterectomy or endometriosis, as well as reimbursements for GnRHa or LNG-IUS during the prior 5 years **(Table 3).** Among MHT initiators, prior gynaecological events and use of selected medications were consistent across age groups, although GnRHa or LNG-IUS use during the previous 5 years was very rare after 60 years **(Additional File 1 eTables 2-9)**. Compared with the reference populations, prior reimbursements for osteoporosis treatments were more frequent among women starting MHT after 60 years.

**Table 3.** Characteristics at index date of MHT initiators and a reference population of non-initiators.

|  | Non-MHT<br>initiators |  | MHT initiators |  |  |  |  |  |
| --- | --- | --- | --- | --- | --- | --- | --- | --- |
|  |  |  | All |  | Starting with<br>unopposed<br>oestrogens |  | Starting with<br>combined therapy |  |
| Cohort, No. | 2,314,672 |  | 578,668 |  | 117,303 |  | 461,365 |  |
| Socio-demographic characteristics |  |  |  |  |  |  |  |  |
| Age, years (median [IQR]) | 52<br>[50 - 55] |  | 52<br>[50 - 55] |  | 52<br>[49 - 56] |  | 52<br>[50- 55] |  |
| Age group, years (n, %) |  |  |  |  |  |  |  |  |
| 45 to 49 | 523,084 | 22.6 | 130,771 | 22.6 | 29,538 | 25.2 | 101,233 | 21.9 |
| 50 to 54 | 1,147,260 | 49.6 | 286,815 | 49.6 | 50,491 | 43.0 | 236,324 | 51.2 |
| 55 to 59 | 473,556 | 20.5 | 118,389 | 20.5 | 19,499 | 16.6 | 98,890 | 21.4 |
| 60 to 64 | 88,772 | 3.8 | 22,193 | 3.8 | 7,120 | 6.1 | 15,073 | 3.3 |
| 65 to 69 | 43,452 | 1.9 | 10,863 | 1.9 | 4,834 | 4.1 | 6,029 | 1.3 |
| 70 to 74 | 20,604 | 0.9 | 5,151 | 0.9 | 2,773 | 2.4 | 2,378 | 0.5 |
| 75 to 79 | 10,080 | 0.4 | 2,520 | 0.4 | 1,565 | 1.3 | 955 | 0.2 |
| 80+ | 7,864 | 0.3 | 1,966 | 0.3 | 1,483 | 1.3 | 483 | 0.1 |
| Complementary health insurance for low-income individuals, (n, %) | 152,127 | 6.6 | 30,263 | 5.2 | 6,868 | 5.9 | 23,395 | 5.1 |
| Outpatient healthcare seeking in the prior 12 months |  |  |  |  |  |  |  |  |
| Number of visits with a gynaecologist, (median [IQR]) | 0 [0 - 1] |  | 1 [1 - 2] |  | 1 [0 - 2] |  | 1 [1 - 2] |  |
| Number of visits with a general practitioner, (median [IQR]) | 3 [0 - 6] |  | 5 [2 - 8] |  | 5 [3 - 9] |  | 5 [2 - 8] |  |
| Number of visits with other healthcare professionals, (median [IQR]) | 2 [0 - 8] |  | 6 [3 - 17] |  | 7 [3 - 19] |  | 6 [3 - 16] |  |
| Mammography in the prior 6 months, (n, %) | 265,926 | 11.5 | 162,343 | 28.1 | 27,898 | 23.8 | 134,445 | 29.1 |

|  | Non-MHT initiators |  | MHT initiators |  |  |  |  |  |
| --- | --- | --- | --- | --- | --- | --- | --- | --- |
|  |  |  | All |  | Starting with unopposed oestrogens |  | Starting with combined therapy |  |
| Cohort, No. | 2,314,672 |  | 578,668 |  | 117,303 |  | 461,365 |  |
| History of gynaecological events in the prior 5 years, (n, %) |  |  |  |  |  |  |  |  |
| Hysterectomy | 42,533 | 1.8 | 34,601 | 6.0 | 27,400 | 23.4 | 7,201 | 1.6 |
| Bilateral oophorectomy | 11,242 | 0.5 | 7,908 | 1.4 | 2,807 | 2.4 | 5,101 | 1.1 |
| Hospitalisation for endometriosis | 20,208 | 0.9 | 16,847 | 2.9 | 10,888 | 9.3 | 5,959 | 1.3 |
| Infertility treatments | 4,516 | 0.2 | 7,282 | 1.3 | 1,127 | 1.0 | 6,155 | 1.3 |
| History of potential MHT contra-indications in the prior 5 years, (n, %) |  |  |  |  |  |  |  |  |
| Breast cancer | 42,035 | 1.8 | 706 | 0.1 | 292 | 0.2 | 414 | 0.1 |
| Ovarian cancer | 2,121 | 0.1 | 516 | 0.1 | 376 | 0.3 | 140 | 0 |
| Uterine cancer | 2,005 | 0.1 | 400 | 0.1 | 318 | 0.3 | 82 | 0 |
| Other cancers | 43,395 | 1.9 | 9,438 | 1.6 | 3,363 | 2.9 | 6,075 | 1.3 |
| Venous thromboembolism | 12,101 | 0.5 | 1,384 | 0.2 | 457 | 0.4 | 927 | 0.2 |
| Cardio neurovascular disease (other than venous thromboembolism) | 59,205 | 2.6 | 10,447 | 1.8 | 3,519 | 3.0 | 6,928 | 1.5 |
| History of other medications in the prior 5 years, (n, %) |  |  |  |  |  |  |  |  |
| Treatment for osteoporosis | 34,747 | 1.5 | 11,298 | 2.0 | 3,300 | 2.8 | 7,998 | 1.7 |
| Gonadotropin releasing hormone agonists | 11,004 | 0.5 | 9,015 | 1.6 | 3,118 | 2.7 | 5,897 | 1.3 |
| Vaginal oestrogens | 244,295 | 10.6 | 155,851 | 26.9 | 35,249 | 30.0 | 120,602 | 26.1 |
| Levonorgestrel-releasing intrauterine system | 134,320 | 5.8 | 45,195 | 7.8 | 16,854 | 14.4 | 28,341 | 6.1 |

Abbreviations: IQR – interquartile range; MHT – menopausal hormone therapy

### Treatment patterns among MHT initiators

Among MHT initiators aged 45 years or older, the median follow-up was 9.0 [IQR 4.9 -12.3] years. The median cumulative treatment duration was 2.1 [IQR 0.6-6.1] years overall. It was stable among women starting treatment between 2011 and 2019, and slightly increased afterwards (2.5 years among 2022 initiators), noting that the median was not reached among women starting treatment after 2023. Treatment duration was longer among MHT initiators aged 50-54 years, among those using transdermal oestrogens or regimens with micronised progesterone or dydrogesterone **(Table 4 and Additional File 1 eTable 10)**. Overall, treatment spanned 2 [IQR 1-4] episodes, with a first interruption after 1 [IQR 0.4-3.5] year, slightly earlier among unopposed oestrogen initiators. Among the 308,093 MHT initiators with at least two treatment episodes, treatment resumed after 0.4 [IQR 0.3-0.7] years, similar across regimens (**Table 4)**.

**Table 4.** Temporal patterns of systemic MHT among initiators: treatment duration and treatment episodes.

| Characteristics | All MHT initiators |  | Starting with unopposed oestrogens |  | Starting with combined therapy |  |
| --- | --- | --- | --- | --- | --- | --- |
| Cohort, No. | 578,668 |  | 117,303 |  | 461,365 |  |
|  | Median | IQR | Median | IQR | Median | IQR |
| Total length of the follow-up (years) | 9.0 | 4.9 – 12.3 | 9.2 | 5.2 – 12.3 | 9.0 | 4.9 – 12.2 |
| Time between the first and the last MHT use (years) | 3.3 | 0.9 – 7.9 | 2.5 | 0.7 – 6.8 | 3.6 | 1.0 – 8.1 |
| Cumulative MHT duration (years) | 2.1 | 0.6 – 6.1 | 1.4 | 0.5 – 4.8 | 2.4 | 0.7 – 6.4 |
| Number of MHT episodes | 2 | 1 - 4 | 2 | 1 - 4 | 2 | 1 - 4 |
| Duration of the first MHT episode (years) | 1.0 | 0.4 – 3.5 | 0.6 | 0.3 – 2.2 | 1.1 | 0.4 – 3.8 |
| Duration of the pause between the first two MHT episodes (years)* | 0.4 | 0.3 - 0.7 | 0.5 | 0.3 - 0.8 | 0.4 | 0.3 - 0.7 |

Abbreviations: IQR – interquartile range; MHT – menopausal hormone therapy \* Among 308,093 incident MHT users for whom at least two exposure episodes were observed

During follow-up, among unopposed oestrogen initiators, 22.0% switched at least once to combined therapy, with combined therapy representing 20.2% of all person-months of MHT use in this group **(Table 5)**. Conversely, among combined therapy initiators, 13.1% switched at least once to unopposed oestrogens, which represented 4.2% of all person-months of treatment in this group. Oestradiol was used quasi-exclusively as the oestrogen component. Transdermal administration was used exclusively by 74.2% of MHT initiators, oral administration by 12.1%, and both routes by 13.7% (rarely concomitantly). Overall, transdermal oestrogens accounted for 81.6% of total person-months of treatment, with gels being the predominant formulation. Among 487,165 MHT initiators receiving combined therapies at least once during follow-up, 62.9% used micronised progesterone exclusively, 9.2% used dydrogesterone, and 21.9% used multiple progestogens **(Table 5)**. Among MHT initiators, 38.6% had at least one reimbursement for vaginal oestrogens during follow-up, 22.1% concomitant with systemic MHT use, and 9.5% in the 30 days around systemic MHT initiation. Gynaecologists initiated the majority of systemic MHTs and remained the primary prescribers during follow-up, followed by general practitioners **(Table 5 and Additional File 1 eTables 11)**.

**Table 5.** Treatment-related characteristics during follow-up among MHT initiators aged 45 years or older.

| Characteristic | All MHT initiators |  | Starting with unopposed oestrogens |  | Starting with combined therapy |  |
| --- | --- | --- | --- | --- | --- | --- |
| Cohort, No. | 578,668 |  | 117,303 |  | 461,365 |  |
| Systemic MHT used during follow-up |  |  |  |  |  |  |
| Treatment regimens |  |  |  |  |  |  |
| Unopposed oestrogens, at least once, n (%) | 177,842 | 30.7 | 117,303 | 100.0 | 60,539 | 13.1 |
| Combined therapy, at least once, n (%) | 487,165 | 84.2 | 25,800 | 22.0 | 461,365 | 100.0 |
| Unopposed oestrogens, % person-months* | 17.4 |  | 79.8 |  | 4.2 |  |
| Combined therapy, % person-months* | 82.7 |  | 20.2 |  | 95.8 |  |
| Molecule of oestrogen, n (%) |  |  |  |  |  |  |
| Oestradiol only | 570,170 | 98.5 | 110,389 | 94.1 | 459,781 | 99.7 |
| Oestradiol and estriol | 2,541 | 0.4 | 1,178 | 1.0 | 1,363 | 0.3 |
| Estriol only | 5,957 | 1.0 | 5,736 | 4.9 | 221 | 0.0 |
| Oestrogen administration routes |  |  |  |  |  |  |
| Transdermal only, n (%) | 429,381 | 74.2 | 90,314 | 77.0 | 339,067 | 73.5 |
| Oral only, n (%) | 70,295 | 12.1 | 11,434 | 9.7 | 58,861 | 12.8 |
| Oral and transdermal, n (%)** | 78,992 | 13.7 | 15,555 | 13.3 | 63,437 | 13.7 |
| Transdermal, % person-months* | 82.7 |  | 85.6 |  | 82.0 |  |
| Oral, % person-months* | 18.0 |  | 15.1 |  | 18.6 |  |
| Oestrogen pharmaceutical forms, n (%) |  |  |  |  |  |  |
| Oral pills only | 78,992 | 13.7 | 15,555 | 13.3 | 63,437 | 13.7 |
| Transdermal patches only | 36,370 | 6.3 | 10,825 | 9.2 | 25,545 | 5.5 |
| Transdermal gels only | 346,722 | 59.9 | 69,164 | 59.0 | 277,558 | 60.2 |
| Oral pills and transdermal patches | 7,505 | 1.3 | 1,333 | 1.1 | 6,172 | 1.3 |
| Oral pills and transdermal gels | 51,352 | 8.9 | 8,058 | 6.9 | 43,294 | 9.4 |
| Transdermal patches and gels | 46,289 | 8.0 | 10,325 | 8.8 | 35,964 | 7.8 |
| Oral pills and transdermal patches and gels | 11,438 | 2.0 | 2,043 | 1.7 | 9,395 | 2.0 |
| Type of progestogens |  |  |  |  |  |  |
| Micronised progesterone or dydrogesterone, n (%) | 407,725 | 70.5 | 21,830 | 18.6 | 385,895 | 83.6 |
| Progestogens other than progesterone/dydrogesterone, n (%) | 37,657 | 6.5 | 2,122 | 1.8 | 35,535 | 7.7 |
| Both, n (%) | 41,783 | 7.2 | 1,848 | 1.6 | 39,935 | 8.7 |
| None, n (%) | 91,503 | 15.8 | 91,503 | 78.0 | 0 | 0.0 |
| Micronised progesterone or dydrogesterone, % person-months* | 76.2 |  | 18.8 |  | 88.4 |  |
| Progestogens other than progesterone/dydrogesterone, % person-months* | 6.4 |  | 1.4 |  | 7.4 |  |
| Molecule of progestogen (among the 487,165 women using combined therapies at least once) |  |  |  |  |  |  |
| Chlormadinone, n (%) | 8,155 | 1.7 | 427 | 1.6 | 7,728 | 1.7 |
| Cyproterone acetate, n (%) | 1,360 | 0.3 | 89 | 0.3 | 1,271 | 0.3 |
| Dienogest, n (%) | 295 | 0.1 | 9 | 0.0 | 286 | 0.1 |
| Dydrogesterone, n (%) | 45,020 | 9.2 | 2,552 | 9.8 | 42,468 | 9.2 |
| Gestodene, n (%) | 153 | 0.0 | 8 | 0.0 | 145 | 0.0 |
| Levonorgestrel, n (%) | 1,938 | 0.4 | 187 | 0.7 | 1,751 | 0.4 |
| Medrogestone, n (%) | 1,343 | 0.3 | 97 | 0.4 | 1,246 | 0.3 |
| Medroxyprogesterone acetate, n (%) | 991 | 0.2 | 66 | 0.3 | 925 | 0.2 |
| Nomegestrol, n (%) | 7,365 | 1.5 | 413 | 1.6 | 6,952 | 1.5 |
| Norethisterone acetate, n (%) | 5,858 | 1.2 | 329 | 1.3 | 5,529 | 1.2 |
| Micronised progesterone, n (%) | 306,539 | 62.9 | 16,687 | 64.2 | 289,852 | 62.8 |
| Promegestone, n (%) | 1,827 | 0.4 | 96 | 0.4 | 1,731 | 0.4 |
| Multiple molecules, n (%) | 106,498 | 21.9 | 5,017 | 19.3 | 101,481 | 22.0 |
| **Gynaecological events during the follow-up, n (%)** |  |  |  |  |  |  |
| Hysterectomy | 12,872 | 2.2 | 2,312 | 2.0 | 10,560 | 2.3 |
| Bilateral oophorectomy | 8,374 | 1.4 | 1,797 | 1.5 | 6,577 | 1.4 |
| Hospitalisation for endometriosis | 7,502 | 1.3 | 1,465 | 1.2 | 6,037 | 1.3 |
| Infertility treatments | 4,660 | 0.8 | 905 | 0.8 | 3,755 | 0.8 |
| **Reimbursements of other medications during the follow-up** |  |  |  |  |  |  |
| Treatments for osteoporosis | 28,763 | 5.0 | 6,207 | 5.3 | 22,556 | 4.9 |
| Levonorgestrel-releasing intrauterine system, n (%) | 4,123 | 0.7 | 2,077 | 1.8 | 2,046 | 0.4 |
| Gonadotropin-releasing hormone agonists, n (%) | 4,860 | 0.8 | 1,231 | 1.0 | 3,629 | 0.8 |
| Vaginal oestrogens, n (%) |  |  |  |  |  |  |
| At least one reimbursement | 223,473 | 38.6 | 46,256 | 39.4 | 177,217 | 38.4 |
| Concomitant with systemic MHT initiation ($\pm 30$ days) | 54,702 | 9.5 | 14,240 | 12.1 | 40,462 | 8.8 |
| At least one reimbursement during the systemic MHT treatment (concomitant use) | 127,719 | 22.1 | 28,231 | 24.1 | 99,488 | 21.6 |
| **Speciality of MHT prescribers** |  |  |  |  |  |  |
| At initiation (missing = 62), n (%) |  |  |  |  |  |  |
| General practitioner (GP) | 194,545 | 33.6 | 48,043 | 41.0 | 146,502 | 31.8 |
| Gynaecologist | 360,769 | 62.3 | 62,765 | 53.5 | 298,004 | 64.6 |
| Other | 23,292 | 4.0 | 6,489 | 5.5 | 16,803 | 3.6 |
| During follow-up, n (%) |  |  |  |  |  |  |
| GP only | 119,846 | 20.7 | 33,024 | 28.2 | 86,822 | 18.8 |
| Gynaecologist only | 167,809 | 29.0 | 29,591 | 25.2 | 138,218 | 30.0 |
| GP & Gynaecologist | 196,549 | 34.0 | 35,906 | 30.6 | 160,643 | 34.8 |
| GP & Gynaecologist & Other | 54,939 | 9.5 | 9,236 | 7.9 | 45,703 | 9.9 |
| GP & Other | 17,508 | 3.0 | 4,782 | 4.1 | 12,726 | 2.8 |
| Gynaecologist & Other | 15,495 | 2.7 | 2,533 | 2.2 | 12,962 | 2.8 |
| Other | 6,522 | 1.1 | 2,231 | 1.9 | 4,291 | 0.9 |

Abbreviations: GP – General practitioner; MHT – Menopausal Hormone therapy;
\* Relative to the total number of months of MHT use (rate per 100 months)
\*\* Simultaneous or sequential intake

Among MHT initiators aged 50-59 years, the median treatment duration was 2.4 [IQR 0.7-6.4] years. Transdermal oestrogens were exclusively used by 77.8% of women, and micronised progesterone by 66.2% MHT initiators receiving combined therapies at least once during follow-up (**Additional File 1 eTables 12-13**).

## Discussion

### Main findings

This study provides a comprehensive overview of systemic MHT use in France between 2011 and 2024. After a prolonged decline, the use of MHT increased again slightly between 2022 and 2024 among women aged 45 to 59 years. However, overall prevalence of MHT treatment remained low, with regimens predominantly combining transdermal oestrogen and micronised progesterone or dydrogesterone. At initiation, which occurred at a median age of 52 years, MHT users had a lower prevalence of prior breast cancer and cardio-neurovascular diseases than non-initiators. Gynaecologists predominantly prescribed treatments. Treatment duration was relatively short (median 2 years), typically spanning two episodes, with an initial interruption after 1 year.

### Interpretation of results and comparison with other studies

Consistent with our findings, two recent French studies have reported a persistent decline in systemic MHT utilisation levels up to 2022: an institutional report using the SNDS database, and a nationwide study using open, aggregated drug-claims data.[39, 40] The institutional report showed increased MHT use after 2022 and increased initiation since 2020 among women aged 45-60 years,[39] while the aggregated drug-claims study reported a continued decline in prevalence until 2023.[40] These reports lack information on discontinuation proportions. Our study shows a recent decrease in MHT discontinuations, which suggests that the recent increase in MHT prevalence among women aged 45 to 59 years after 2022 results from both increased treatment initiation and decreased treatment discontinuation. The lower proportion of initiation in 2020 likely reflects COVID-19-related disruptions in health care access rather than a true reversal in prescribing practices. These findings may reflect renewed clinical interest in France in systemic MHT for managing climacteric symptoms and preventing osteoporosis.[41]

Overall, systemic MHT prescribing patterns in France appear broadly consistent with current guidelines. In France, since 2003, guidelines have restricted systemic MHT indication to climacteric symptoms substantially affecting quality of life and to osteoporosis prevention in women at risk of osteoporotic fracture who cannot use first-line treatments. Current recommendations call for the lowest effective dose and duration, tailored to each woman’s risk-benefit profile and reassessed regularly.[41–43] Among notable and original findings of our study, the short treatment durations, with a first interruption after 1 year, possibly reflect regular treatment reassessment. The median age at systemic MHT initiation (52 years) was close to the age at natural menopause in France (51 years), indicating that MHT was generally prescribed for climacteric symptoms occurring near menopause onset. The more frequent history of osteoporosis treatments among women starting systemic MHT after 60 years suggests that MHT may have been prescribed for osteoporosis prevention in older age groups. The lower prevalence of previous breast cancer and cardio-neurovascular events, both potential contraindications to systemic MHT prescribing, among MHT initiators compared with non-initiators also aligns with appropriate prescribing.

Despite a median treatment duration of 2 years, a quarter of initiators were treated for less than 6 months, and another quarter for over 5.7 years. The wide distribution, along with differential duration patterns according to age, regimens, and oestrogen administration routes suggest substantial heterogeneity in treatment trajectories, including persistent climacteric symptoms requiring prolonged treatment in some women, and use of systemic oestrogens for other indications in others. Earlier French studies reported longer treatment durations before the WHI (3.5-8.3 years), followed by a marked decline thereafter.[19, 20] Treatment duration remained stable in our cohort through 2019 before increasing slightly. A recent French institutional report estimated a mean treatment duration of 3.7 years (±3 years) between 2012 and 2024.[39] However, direct comparison is limited by different metrics (mean versus median) and episode identification methods (fixed 6-months intervals versus hypothetical daily dosage versus).

Our results confirm the continued predominance in France of transdermal oestrogen combined with micronised progesterone or dydrogesterone, regimens generally considered to have a more favourable safety profile. Transdermal oestrogen has been associated with a lower risk of VTE than oral oestrogen.[44] Micronised progesterone and dydrogesterone have been associated with a more favourable cardiometabolic profile and lower breast cancer risk than other progestogens.[45] These results extend long-standing prescribing patterns in France, where transdermal oestrogen predominated (58.8-74.5%) and unopposed oestrogen use was limited (12.4-23.2%) since before the WHI publication.[20, 21] The use of micronised progesterone as the progestogen component increased over time, from 18-25% before 2000 to 68% by 2003-2007.[19–21] Consistent with these trends, only 16% of combined therapy initiators used progestogens other than progesterone or dydrogesterone between 2011 and 2024 in our study. Our findings are similar to those identified in the institutional report among women starting treatment in 2024.[39]

Among systemic MHT initiators, nearly 80% of women started and generally remained on therapies combining oestrogen and progestogen. The rare switches to unopposed oestrogen might reflect hysterectomies during follow-up or isolated regimen misclassification. In contrast, nearly one-quarter of women starting with unopposed oestrogen later used combined therapy. This may indicate occasional prescribing of unopposed oestrogen to non-hysterectomised women. Alternatively, some women may have received endometrial protection through an LNG-IUS at treatment initiation before subsequently switching to conventional progestogens, a practice recommended in some countries, but not officially endorsed in France.[38, 45, 46] Consistent with this, prior LNG-IUS reimbursement was more common among unopposed oestrogen initiators aged 45-59 years than among combined therapy initiators. Additionally, prior hospitalisation for severe endometriosis was more frequent among women aged 45-59 years starting with unopposed oestrogens, suggesting that, in some cases, unopposed oestrogen might have been prescribed for indications other than menopausal symptoms.

### Strengths and limitations

Strengths of our study include the use of a large, nationwide, individual-level database, allowing a detailed description of MHT utilisation patterns in France, including treatment regimens, duration, and treatment episodes, across fine-grained age groups. Comparing the characteristics of systemic MHT initiators with those of a reference population of non-initiators allowed interpreting observed prescribing patterns and inform on their consistency with prescription guidelines.

Our study also presents several limitations, specific to the use of routinely collected claims data. First, the prescribed regimen, dose, and treatment duration of systemic MHT are unavailable in the SNDS. Thus, despite pairing oestrogen and progestogen reimbursements less than 3 months apart, residual MHT regimen misclassification may remain when products were purchased separately. However, both products were dispensed concomitantly in 92% of combined regimens involving separate products, suggesting limited impact. Second, the menopause status, type, age at onset, or treatment indication are not available in the SNDS, preventing direct identification of postmenopausal women. However, 5-year age-stratified analyses beyond 51 years (the median age of natural menopause in France [38]) provide a good approximation of postmenopausal MHT use. Some younger initiators may have received systemic oestrogens for other indications, such as endometriosis or as add-back therapy. Third, non-reimbursed MHT drugs, including tibolone (ATC5 G03CX01), are not captured in the SNDS, but these products account for only 10% of total MHT sales.[40] Finally, healthcare data pre-2006 are unavailable in the SNDS, likely resulting in under-detection of the history of health events at the index date. However, using an equal 5-year look-back period for all women should have limited differential misclassifications.

### Conclusion and Perspectives

Systemic MHT use in France remained low between 2011 and 2024, with a prolonged decline followed by a modest and recent increase in treatment initiation among women aged 45 to 64 years. Treatment consisted predominantly of MHT regimens combining transdermal oestradiol with micronised progesterone or dydrogesterone, prescribed for relatively short durations. This study provides a comprehensive overview of real-world use of systemic MHT in France, allowing public health authorities to monitor alignment between prescribing patterns and recommendations and evaluate future changes in MHT utilisation.

## Data Availability

The datasets generated and or analysed during the current study are not publicly available due to privacy issues but additional results and aggregated findings are available from the corresponding author upon reasonable request.

## Additional Files

***Additional_File_1.docx: Supplementary eTables 1-13 and eFigures 1-8***

eTable 1. Codes for identifying medical events in the SNDS database

eFigure 1. Prevalence of systemic MHT between 2011 and 2024, by oestrogen administration route and MHT regimen

eFigure 2. Proportion of systemic MHT initiators between 2011 and 2024, by oestrogen administration route and MHT regimen

eFigure 3. Proportion of systemic MHT discontinuers between 2011 and 2024, by oestrogen administration route and age group

eFigure 4. Proportion of systemic MHT discontinuers between 2011 and 2024, by MHT regimen and age group

eFigure 5. Age at systemic MHT initiation, according to the MHT regimen at initiation

eTable 2. History of hysterectomy within 5 years before the index date, by age at index date

eTable 3. History of oophorectomy within 5 years before the index date, by age at index date

eTable 4. Osteoporosis treatment reimbursement within 5 years before the index date, by age at index date

eTable 5. Gonadotropin-releasing hormone agonists reimbursements within 5 years before the index date, by age at index date

eTable 6. Vaginal oestrogens reimbursements within 5 years before the index date, by age at index date

eTable 7. Levonorgestrel intra-uterine system reimbursements within 5 years before the index date, by age at index date

eTable 8. Hospitalisation for endometriosis within 5 years before the index date, by age at index date

eTable 9. Infertility treatments within 5 years before the index date, by age at index date Treatment patterns among MHT initiators aged 45 years or older

eTable 10. Cumulative duration of systemic MHT by age, initiation year, oestrogen administration route, and regimen at initiation

eTable 11. Prescriber speciality of all systemic oestrogen reimbursements during the follow-up

eFigure 6. Prevalence, initiation and discontinuation of systemic MHT among women aged 50-59 years

eFigure 7. Prevalence of systemic MHT among women aged 50-59 years, by oestrogen administration route and MHT regimen

eFigure 8. Proportion of systemic MHT initiators among women aged 50-59 years, by oestrogen administration route and MHT regimen

eTable 12. Temporal patterns of systemic MHT among initiators aged 50-59 years: treatment duration and treatment episodes

eTable 13. Treatment-related characteristics during follow-up among MHT initiators aged 50-59 years

## Declarations

### Ethics approval and consent to participate

In accordance with national regulations (Decree no. 2021–848 of June 29, 2021), researchers at Inserm (French National Institute of Health and Medical Research) have a statutory access to data from the SNDS database. The research protocol of the current study is compliant with the internal control process set up by the Inserm SNDS Service Center, in conjunction with the establishment’s data protection delegate (DPO); Internal compliance approval was granted on 20/02/2024. Informed consent from individual patients was not required. In compliance with the legal requirement for transparency towards the public regarding the use of the SNDS, as established by the French Public Health Code, our study was registered in the Health Data Hub public registry on July 31, 2025 (https://www.health-data-hub.fr/projets/description-de-lutilisation-des-traitements-hormonaux-de-la-menopause-dans-la-population).

### Consent for publication

Not applicable

### Availability of data and materials

The datasets generated and/or analysed during the current study are not publicly available due to privacy issues but additional results and aggregated findings are available from the corresponding author upon reasonable request.

### Authors’ contributions

AMV conceived and designed the study, performed the statistical analysis, interpreted the data, drafted, and critically revised the manuscript, PGS, YZ, MC, AF, JB, LM, VV interpreted the data and critically revised the manuscript, AF designed the study, provided supervision, interpreted the data, drafted, and critically revised the manuscript, AT obtained funding, conceived and designed the study, provided supervision, interpreted the data, and critically revised the manuscript. AMV is the guarantor of the study and data integrity. All authors gave approval for the final version of the manuscript and agree to be accountable for all aspects of the work.

## Acknowledgements

Not applicable

## Abbreviations

ATC: Anatomical Therapeutic Chemical
GnRHa: Gonadotropin-releasing hormone agonists
ICD-10: International Classification of Diseases, 10th Edition
GP: General practitioner
LNG-IUS: Levonorgestrel-releasing intrauterine system
MHT: Menopausal hormone therapy
SNDS: Système National des Données de Santé
VTE: Venous thromboembolism
WHI: Women Health Initiative

