## Additional File 1 for "Use of systemic menopausal hormone therapy among French women: a descriptive nationwide study over the 2011 to 2024 period"

**Additional eData Files**

### Codes for identifying medical events in the SNDS database

#### eTable 1. Codes for identifying medical events in the SNDS database

| **Codes** | **Label** |
| --- | --- |
| **Screening mammography** | |
| **CCAM* codes** |  |
| QEQK001 | Bilateral mammography |
| QEQK004 | Screening mammography |
| **Hysterectomy** | |
| **CCAM* codes** |  |
| JKFA021 | Total hysterectomy with unilateral or bilateral adnexectomy and anterior or posterior colpopexy, via vaginal approach |
| JKFA018 | Total hysterectomy, by laparoscopy and vaginal approach |
| JKFA015 | Total hysterectomy, by laparotomy |
| JKFA024 | Subtotal hysterectomy, by laparotomy |
| JKFA025 | Total hysterectomy with anterior or posterior colpopexy, by vaginal approach |
| JKFA028 | Total hysterectomy with unilateral or bilateral adnexectomy, via laparotomy |
| JKFA004 | Total hysterectomy with unilateral or bilateral adnexectomy and posterior suspension of the vaginal vault, via laparotomy |
| JKFA014 | Subtotal hysterectomy with posterior suspension of the cervix [colposuspension], by laparotomy |
| JKFA013 | Total hysterectomy with posterior suspension of the vaginal vault, by laparotomy |
| JKFA002 | Total hysterectomy with anterior and posterior colpopherinoplasty, by vaginal approach |
| JKFA005 | Total hysterectomy with unilateral or bilateral adnexectomy, vaginal approach |
| JKFA001 | Subtotal hysterectomy with unilateral or bilateral adnexectomy and posterior suspension of the cervix [colposuspension], by laparotomy |
| JKFA006 | Total hysterectomy with unilateral or bilateral adnexectomy, via laparoscopy and vaginal approach |
| JKFA012 | Subtotal hysterectomy with posterior suspension of the cervix [colposuspension] and indirect cervicocystopexy to the pectineal ligament [Cooper's ligament], via laparotomy |
| JKFA007 | Total hysterectomy with unilateral or bilateral adnexectomy and anterior and posterior colpopexy, via vaginal approach |
| JKFA029 | Subtotal hysterectomy with unilateral or bilateral adnexectomy, posterior suspension of the cervix [colposuspension] and indirect cervicocystopexy to the pectineal ligament [Cooper's ligament], via laparotomy |
| JKFA026 | Total hysterectomy, via vaginal approach |
| JKFC002 | Subtotal hysterectomy, via laparoscopy |
| JKFC003 | Total hysterectomy with unilateral or bilateral adnexectomy, via laparoscopy |
| JKFC005 | Total hysterectomy, via laparoscopy |
| JKFC006 | Subtotal hysterectomy with unilateral or bilateral adnexectomy, by laparoscopy |
| JKFA032 | Subtotal hysterectomy with unilateral or bilateral adnexectomy, by laparotomy |
| JNFA001 | Hysterectomy for obstetric complications, by laparotomy |
| JKFA023 | Total colpohysterectomy extended to the parametrium, via vaginal approach |
| JKFA027 | Total colpohysterectomy extended to the parametrium, via laparotomy |
| JKFA020 | Total colpohysterectomy extended to the parametrium, via laparoscopy and vaginal approach |
| **Bilateral oophorectomy** | |
| **CCAM* codes** |  |
| JJFA005 | Bilateral oophorectomy, by laparotomy |
| JJFC009 | Bilateral oophorectomy, by laparoscopy |
| JJFA050 | Salpingo-oophorectomy [adnexectomy], by vaginal approach |
| JJFC010 | Salpingo-oophorectomy [adnexectomy], by laparoscopy |
| JJFA004 | Salpingo-oophorectomy [adnexectomy], by laparotomy |
| **Infertility treatments** | |
| **CCAM* codes** |  |
| JJFC011 | Oocyte retrieval by coelioscopy |
| JJFJ001 | Transvaginal oocyte retrieval under ultrasound monitoring |
| JSEC001 | Tubal embryo transfer by coelioscopy |
| JSED001 | Transvaginal intrauterine embryo transfer |
| JSLD001 | Intrauterine artificial insemination |
| JSLD002 | Intracervical artificial insemination |
| YYYY032 | Ovulation induction using gonadotropins followed by artificial insemination or in vitro fertilization |
| **ATC5 codes** |  |
| G03GA01 | Chorionic gonadotropin |
| G03GA02 | Human chorionic gonadotropin |
| G03GA04 | Urofollitropin |
| G03GA05 | Follitropin alpha |
| G03GA06 | Follitropin beta |
| G03GA07 | Lutropin alpha |
| G03GA08 | Choriogonadotropin alpha |
| G03GB02 | Clomifene citrate |
| H01CA01 | Gonadorelin |
| H01CA02 | Nafarelin |
| H01CC01 | Ganirelix acetate |
| H01CC02 | Cetrorelix acetate |
| **GHM codes**** |  |
| 13C16J | Oocyte retrieval, in ambulatory setting |
| **ICD-10 codes** |  |
| Z5280 | Donor or retrieval of oocyte or ovarian tissue |
| **Severe endometriosis** | |
| **ICD-10 codes** |  |
| N80 | Endometriosis |
| **First-line treatments for osteoporosis** | |
| **ATC5 codes** |  |
| M05BA01 | etidronic acid |
| M05BA02 | clodronic acid |
| M05BA03 | pamidronic acid |
| M05BA04 | alendronic acid |
| M05BA05 | tiludronic acid |
| M05BA06 | ibandronic acid |
| M05BA07 | risedronic acid |
| M05BA08 | zoledronic acid |
| G03XC01 | raloxifene |
| M05BX06 | romosozumab |
| H05AA02 | teriparatide |
| M05BX04 | denosumab |

** French classification of medical acts (Classification Commune des Actes Médicaux, CCAM)*

*** French classification of diagnosis-related groups* *(Groupe Homogène des Malades, GHM)*

### Annual prevalence, initiation and discontinuation of systemic MHT – women aged 45 years or older

#### eFigure 1. Prevalence of systemic MHT between 2011 and 2024, by oestrogen administration route and MHT regimen

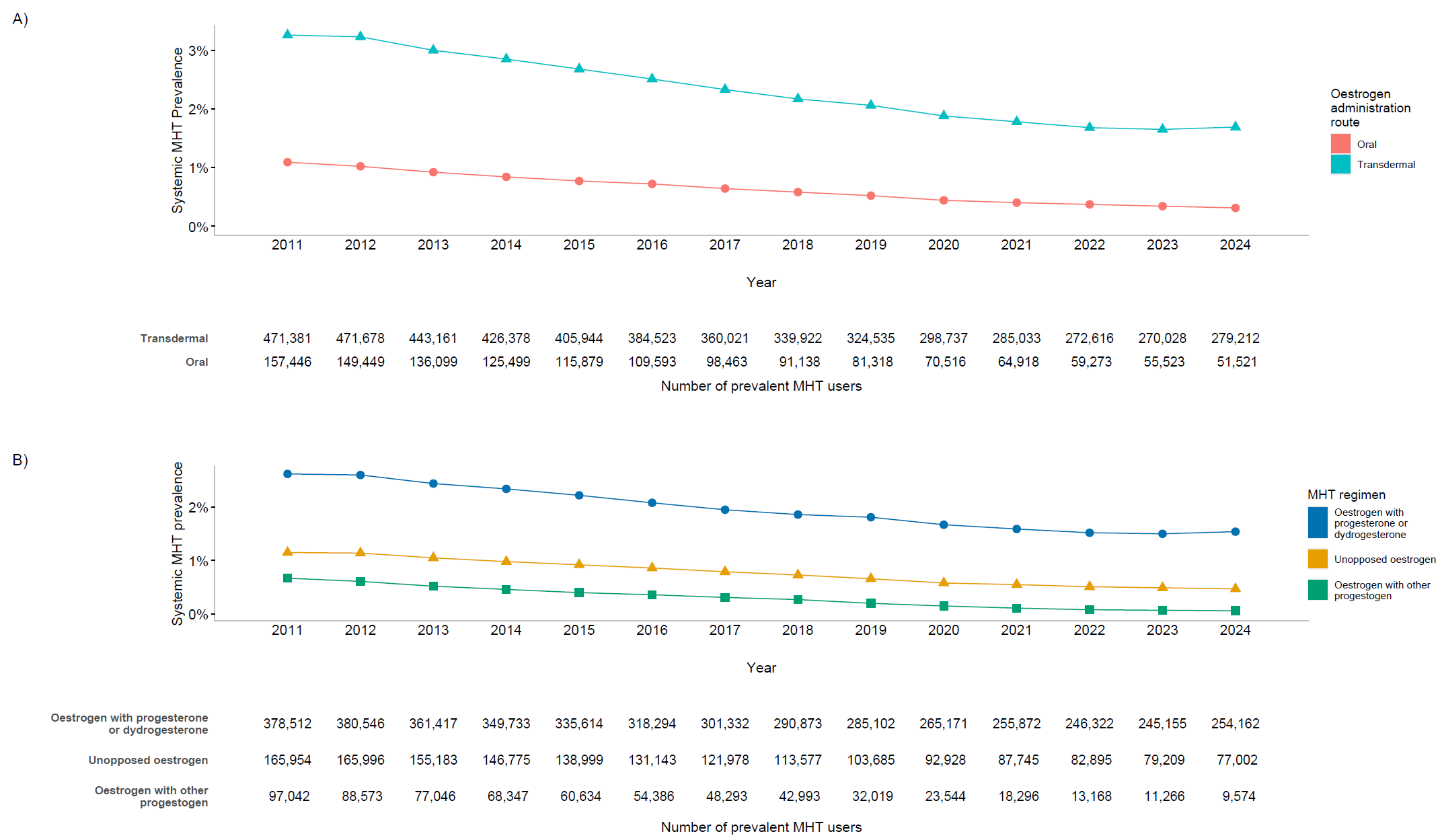

#### eFigure 2. Proportion of systemic MHT initiators between 2011 and 2024, by oestrogen administration route and MHT regimen

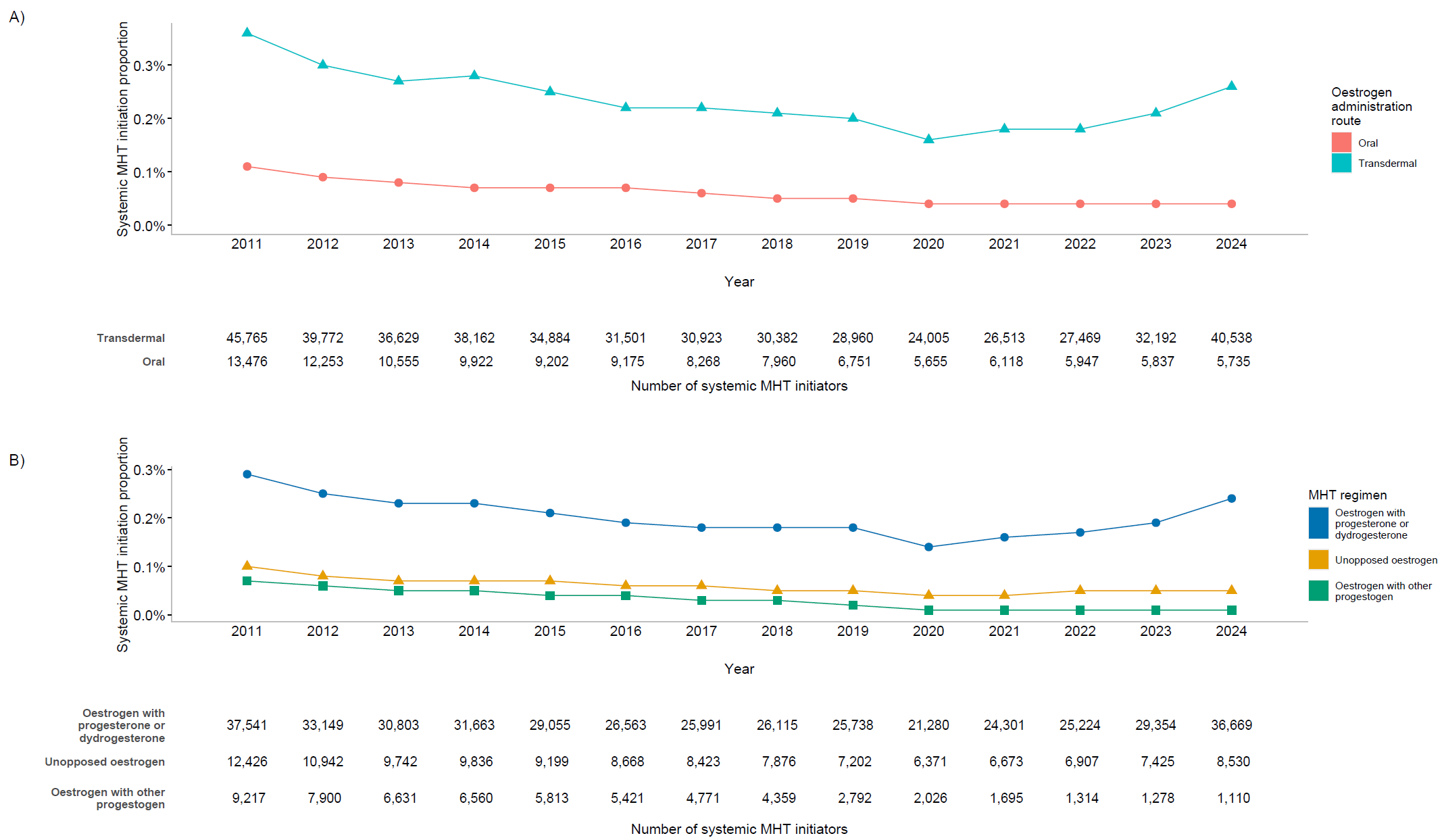

#### eFigure 3. Proportion of systemic MHT discontinuers between 2011 and 2024, by oestrogen administration route and age group

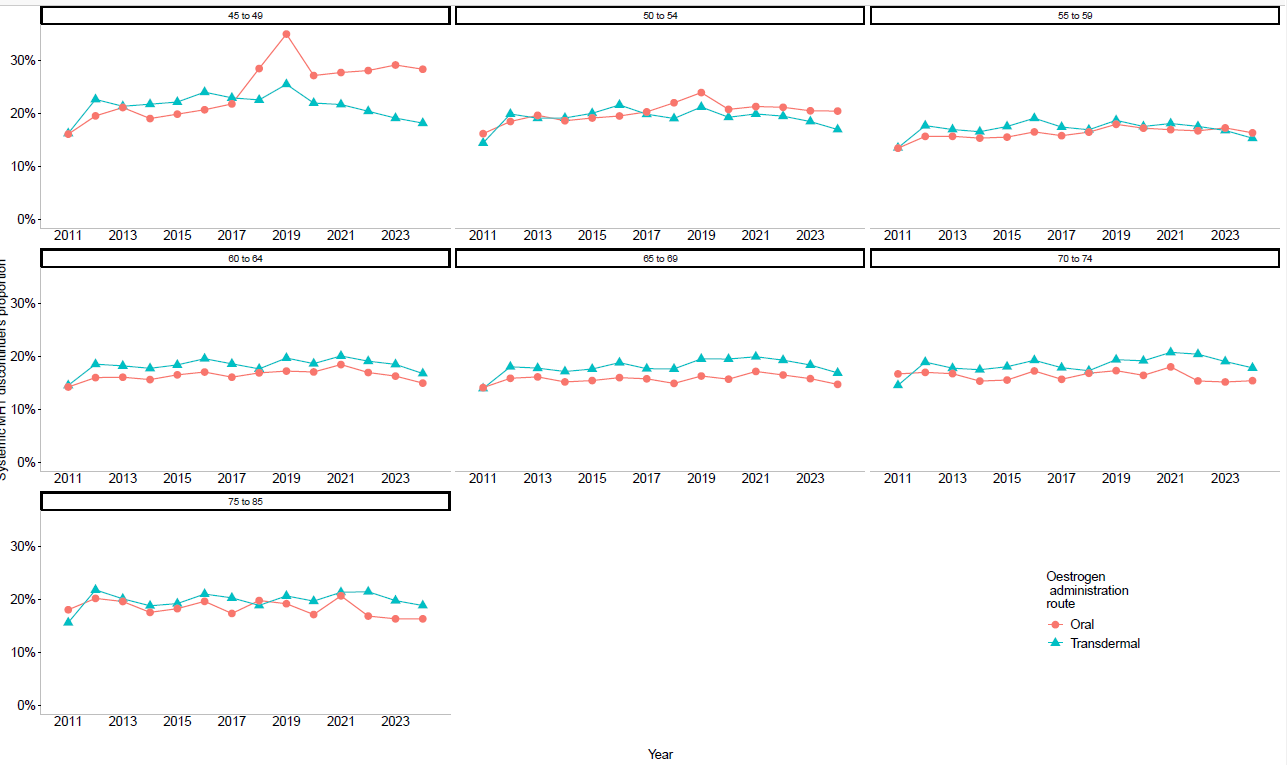

#### eFigure 4. Proportion of systemic MHT discontinuers between 2011 and 2024, by MHT regimen and age group

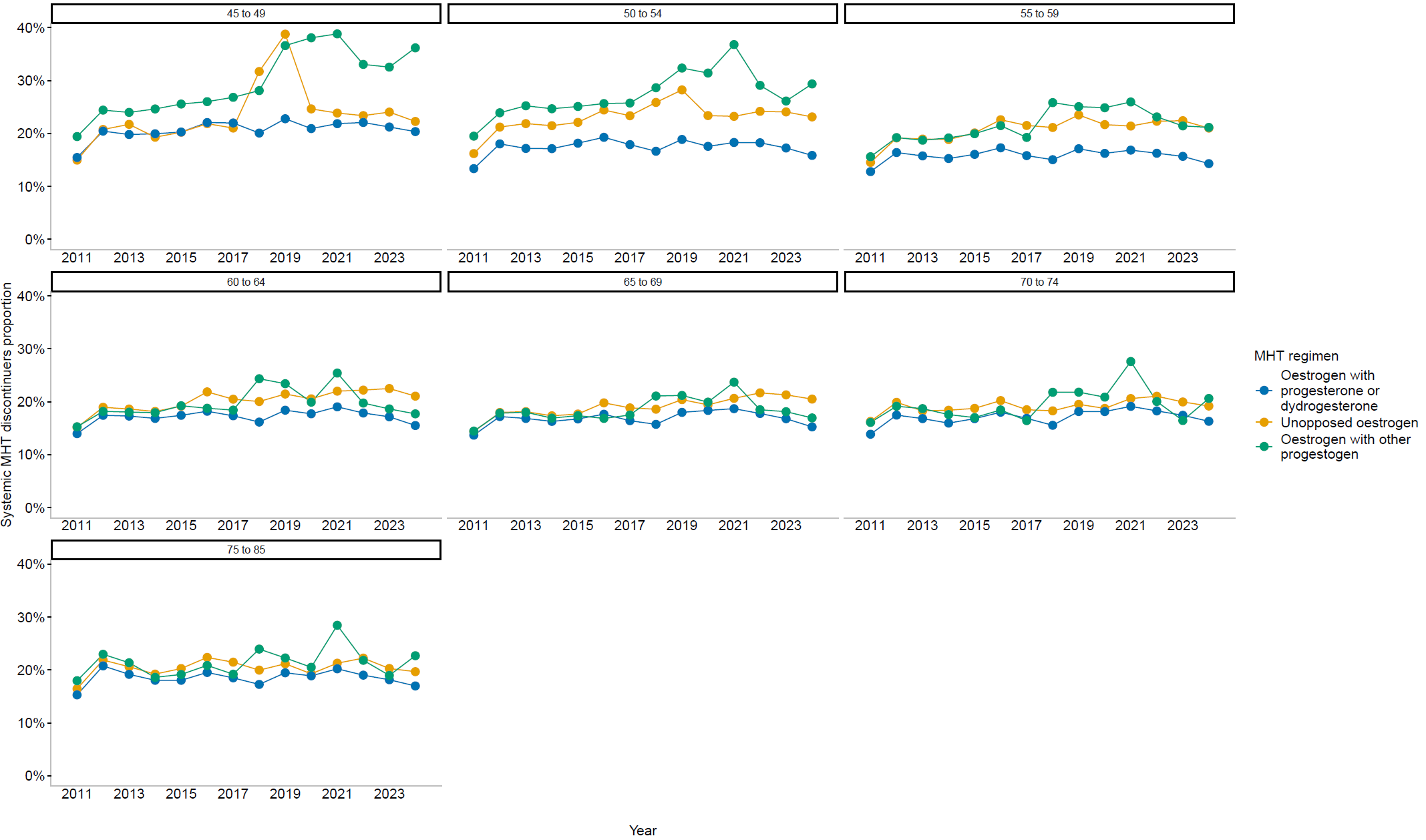

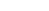

### Characteristics of MHT initiators aged 45 years or older at the index date

#### eFigure 5. Age at systemic MHT initiation, according to the MHT regimen at initiation

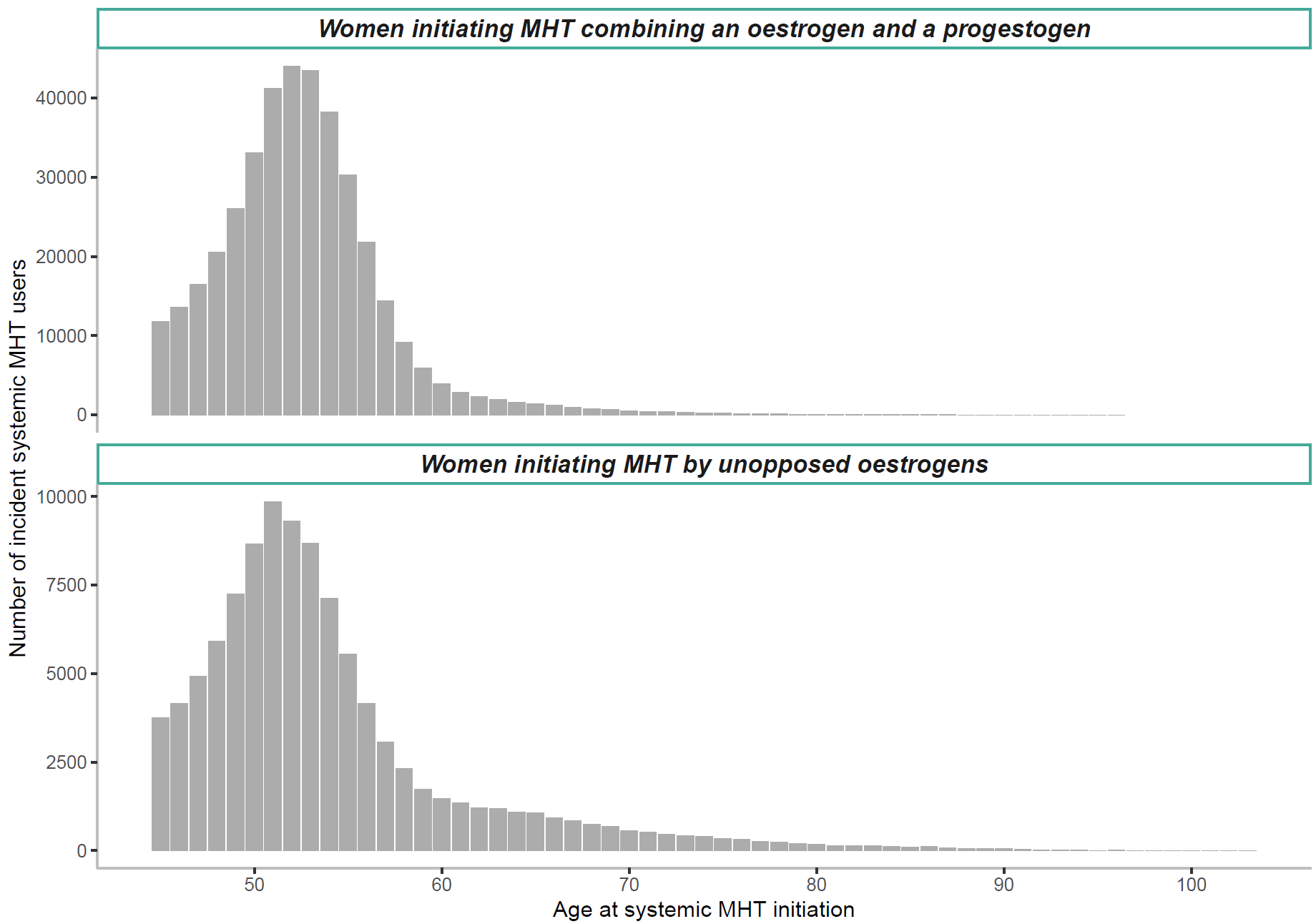

#### eTable 2. History of hysterectomy within 5 years before the index date, by age at index date

|  |  | **Non-MHT initiators** | | |  | **MHT initiators** | | |  | **Starting with unopposed oestrogens** | | |  | **Starting with combined therapy** | | |
| --- | --- | --- | --- | --- | --- | --- | --- | --- | --- | --- | --- | --- | --- | --- | --- | --- |
| **Age group** |  | **N** | **n** | **%** |  | **N** | **n** | **%** |  | **N** | **n** | **%** |  | **N** | **n** | **%** |
| 45 to 49 |  | 523,084 | 11,327 | 2.2 |  | 130,771 | 10,958 | 8.4 |  | 29,538 | 8,421 | 28.5 |  | 101,233 | 2,537 | 2.5 |
| 50 to 54 |  | 1,147,260 | 23,884 | 2.1 |  | 286,815 | 18,402 | 6.4 |  | 50,491 | 14,641 | 29.0 |  | 236,324 | 3,761 | 1.6 |
| 55 to 59 |  | 473,556 | 5,817 | 1.2 |  | 118,389 | 4,355 | 3.7 |  | 19,499 | 3,570 | 18.3 |  | 98,890 | 785 | 0.8 |
| 60 to 64 |  | 88,772 | 774 | 0.9 |  | 22,193 | 484 | 2.2 |  | 7,120 | 418 | 5.9 |  | 15,073 | 66 | 0.4 |
| 65 to 69 |  | 43,452 | 404 | 0.9 |  | 10,863 | 222 | 2.0 |  | 4,834 | 189 | 3.9 |  | 6,029 | 33 | 0.5 |
| 70 to 74 |  | 20,604 | 194 | 0.9 |  | 5,151 | 98 | 1.9 |  | 2,773 | 87 | 3.1 |  | 2,378 | 11 | 0.5 |
| 75 to 79 |  | 10,080 | 81 | 0.8 |  | 2,520 | 53 | 2.1 |  | 1,565 | 46 | 2.9 |  | 955 | 7 | 0.7 |
| 80+ |  | 7,864 | 52 | 0.7 |  | 1,966 | 29 | 1.5 |  | 1,483 | 28 | 1.9 |  | 483 | 1 | 0.2 |

*Abbreviations: MHT – Menopausal Hormone therapy*

#### eTable 3. History of oophorectomy within 5 years before the index date, by age at index date

|  |  | **Non-MHT initiators** | | |  | **MHT initiators** | | |  | **Starting with unopposed oestrogens** | | |  | **Starting with combined therapy** | | |
| --- | --- | --- | --- | --- | --- | --- | --- | --- | --- | --- | --- | --- | --- | --- | --- | --- |
| **Age group** |  | **N** | **n** | **%** |  | **N** | **n** | **%** |  | **N** | **n** | **%** |  | **N** | **n** | **%** |
| 45 to 49 |  | 523,084 | 1,905 | 0.4 |  | 130,771 | 2,741 | 2.1 |  | 29,538 | 1,154 | 3.9 |  | 101,233 | 1,587 | 1.6 |
| 50 to 54 |  | 1,147,260 | 5,809 | 0.5 |  | 286,815 | 3,592 | 1.3 |  | 50,491 | 1,138 | 2.3 |  | 236,324 | 2,454 | 1.0 |
| 55 to 59 |  | 473,556 | 2,634 | 0.6 |  | 118,389 | 1,161 | 1.0 |  | 19,499 | 312 | 1.6 |  | 98,890 | 849 | 0.9 |
| 60 to 64 |  | 88,772 | 508 | 0.6 |  | 22,193 | 227 | 1.0 |  | 7,120 | 97 | 1.4 |  | 15,073 | 130 | 0.9 |
| 65 to 69 |  | 43,452 | 229 | 0.5 |  | 10,863 | 105 | 1.0 |  | 4,834 | 55 | 1.1 |  | 6,029 | 50 | 0.8 |
| 70 to 74 |  | 20,604 | 109 | 0.5 |  | 5,151 | 51 | 1.0 |  | 2,773 | 33 | 1.2 |  | 2,378 | 18 | 0.8 |
| 75 to 79 |  | 10,080 | 35 | 0.3 |  | 2,520 | 27 | 1.1 |  | 1,565 | 15 | 1.0 |  | 955 | 12 | 1.3 |
| 80+ |  | 7,864 | 13 | 0.2 |  | 1,966 | 4 | 0.2 |  | 1,483 | 3 | 0.2 |  | 483 | 1 | 0.2 |

*Abbreviations: MHT – Menopausal Hormone therapy*

#### eTable 4. Osteoporosis treatment reimbursement within 5 years before the index date, by age at index date

|  |  | **Non-MHT initiators** | | |  | **MHT initiators** | | |  | **Starting with unopposed oestrogens** | | |  | **Starting with combined therapy** | | |
| --- | --- | --- | --- | --- | --- | --- | --- | --- | --- | --- | --- | --- | --- | --- | --- | --- |
| **Age group** |  | **N** | **n** | **%** |  | **N** | **n** | **%** |  | **N** | **n** | **%** |  | **N** | **n** | **%** |
| 45 to 49 |  | 523,084 | 1,698 | 0.3 |  | 130,771 | 916 | 0.7 |  | 29,538 | 178 | 0.6 |  | 101,233 | 738 | 0.7 |
| 50 to 54 |  | 1,147,260 | 9,865 | 0.9 |  | 286,815 | 2,749 | 1.0 |  | 50,491 | 438 | 0.9 |  | 236,324 | 2,311 | 1.0 |
| 55 to 59 |  | 473,556 | 10,784 | 2.3 |  | 118,389 | 3,060 | 2.6 |  | 19,499 | 574 | 2.9 |  | 98,890 | 2,486 | 2.5 |
| 60 to 64 |  | 88,772 | 4,931 | 5.6 |  | 22,193 | 1,819 | 8.2 |  | 7,120 | 609 | 8.6 |  | 15,073 | 1,210 | 8.0 |
| 65 to 69 |  | 43,452 | 3,581 | 8.2 |  | 10,863 | 1,313 | 12.1 |  | 4,834 | 609 | 12.6 |  | 6,029 | 704 | 11.7 |
| 70 to 74 |  | 20,604 | 1,965 | 9.5 |  | 5,151 | 726 | 14.1 |  | 2,773 | 402 | 14.5 |  | 2,378 | 324 | 13.6 |
| 75 to 79 |  | 10,080 | 1,039 | 10.3 |  | 2,520 | 382 | 15.2 |  | 1,565 | 237 | 15.1 |  | 955 | 145 | 15.2 |
| 80+ |  | 7,864 | 884 | 11.2 |  | 1,966 | 333 | 16.9 |  | 1,483 | 253 | 17.1 |  | 483 | 80 | 16.6 |

*Abbreviations: MHT – Menopausal Hormone therapy*

#### eTable 5. [Gonadotropin-releasing hormone agonists](https://atcddd.fhi.no/atc_ddd_index/?code=L02AE&showdescription=no) reimbursements within 5 years before the index date, by age at index date

|  |  | **Non-MHT initiators** | | |  | **MHT initiators** | | |  | **Starting with unopposed oestrogens** | | |  | **Starting with combined therapy** | | |
| --- | --- | --- | --- | --- | --- | --- | --- | --- | --- | --- | --- | --- | --- | --- | --- | --- |
| **Age group** |  | **N** | **n** | **%** |  | **N** | **n** | **%** |  | **N** | **n** | **%** |  | **N** | **n** | **%** |
| 45 to 49 |  | 523,084 | 3,443 | 0.7 |  | 130,771 | 5,614 | 4.3 |  | 29,538 | 1,557 | 5.3 |  | 101,233 | 4,057 | 4.0 |
| 50 to 54 |  | 1,147,260 | 6,202 | 0.5 |  | 286,815 | 2,748 | 1.0 |  | 50,491 | 1,284 | 2.5 |  | 236,324 | 1,464 | 0.6 |
| 55 to 59 |  | 473,556 | 1,316 | 0.3 |  | 118,389 | 611 | 0.5 |  | 19,499 | 251 | 1.3 |  | 98,890 | 360 | 0.4 |
| 60 to 64 |  | 88,772 | 34 | <0.1 |  | 22,193 | 29 | 0.1 |  | 7,120 | 17 | 0.2 |  | 15,073 | 12 | 0.1 |
| 65 to 69 |  | 43,452 | 4 | <0.1 |  | 10,863 | 11 | 0.1 |  | 4,834 | 7 | 0.1 |  | 6,029 | 4 | 0.1 |
| 70 to 74 |  | 20,604 | 3 | <0.1 |  | 5,151 | 1 | <0.1 |  | 2,773 | 1 | <0.1 |  | 2,378 | 0 | <0.1 |
| 75 to 79 |  | 10,080 | 1 | <0.1 |  | 2,520 | 1 | <0.1 |  | 1,565 | 1 | 0.1 |  | 955 | 0 | <0.1 |
| 80+ |  | 7,864 | 1 | <0.1 |  | 1,966 | 0 | <0.1 |  | 1,483 | 0 | <0.1 |  | 483 | 0 | <0.1 |

*Abbreviations: MHT – Menopausal Hormone therapy*

#### eTable 6. Vaginal oestrogens reimbursements within 5 years before the index date, by age at index date

|  |  | **Non-MHT initiators** | | |  | **MHT initiators** | | |  | **Starting with unopposed oestrogens** | | |  | **Starting with combined therapy** | | |
| --- | --- | --- | --- | --- | --- | --- | --- | --- | --- | --- | --- | --- | --- | --- | --- | --- |
| **Age group** |  | **N** | **n** | **%** |  | **N** | **n** | **%** |  | **N** | **n** | **%** |  | **N** | **n** | **%** |
| 45 to 49 |  | 523,084 | 35,670 | 6.8 |  | 130,771 | 26,374 | 20.2 |  | 29,538 | 5,961 | 20.2 |  | 101,233 | 20,413 | 20.2 |
| 50 to 54 |  | 1,147,260 | 110,759 | 9.7 |  | 286,815 | 70,336 | 24.5 |  | 50,491 | 12,708 | 25.2 |  | 236,324 | 57,628 | 24.4 |
| 55 to 59 |  | 473,556 | 67,489 | 14.3 |  | 118,389 | 39,206 | 33.1 |  | 19,499 | 7,144 | 36.6 |  | 98,890 | 32,062 | 32.4 |
| 60 to 64 |  | 88,772 | 16,319 | 18.4 |  | 22,193 | 9,945 | 44.8 |  | 7,120 | 3,512 | 49.3 |  | 15,073 | 6,433 | 42.7 |
| 65 to 69 |  | 43,452 | 8,223 | 18.9 |  | 10,863 | 5,214 | 48.0 |  | 4,834 | 2,614 | 54.1 |  | 6,029 | 2,600 | 43.1 |
| 70 to 74 |  | 20,604 | 3,523 | 17.1 |  | 5,151 | 2,471 | 48.0 |  | 2,773 | 1,530 | 55.2 |  | 2,378 | 941 | 39.6 |
| 75 to 79 |  | 10,080 | 1,503 | 14.9 |  | 2,520 | 1,256 | 49.8 |  | 1,565 | 890 | 56.9 |  | 955 | 366 | 38.3 |
| 80+ |  | 7,864 | 809 | 10.3 |  | 1,966 | 1,049 | 53.4 |  | 1,483 | 890 | 60.0 |  | 483 | 159 | 32.9 |

*Abbreviations: MHT – Menopausal Hormone therapy*

#### eTable 7. Levonorgestrel intra-uterine system reimbursements within 5 years before the index date, by age at index date

|  |  | **Non-MHT initiators** | | |  | **MHT initiators** | | |  | **Starting with unopposed oestrogens** | | |  | **Starting with combined therapy** | | |
| --- | --- | --- | --- | --- | --- | --- | --- | --- | --- | --- | --- | --- | --- | --- | --- | --- |
| **Age group** |  | **N** | **n** | **%** |  | **N** | **n** | **%** |  | **N** | **n** | **%** |  | **N** | **n** | **%** |
| 45 to 49 |  | 523,084 | 65,411 | 12.5 |  | 130,771 | 16,575 | 12.7 |  | 29,538 | 6,698 | 22.7 |  | 101,233 | 9,877 | 9.8 |
| 50 to 54 |  | 1,147,260 | 64,697 | 5.6 |  | 286,815 | 24,893 | 8.7 |  | 50,491 | 9,042 | 17.9 |  | 236,324 | 15,851 | 6.7 |
| 55 to 59 |  | 473,556 | 4,191 | 0.9 |  | 118,389 | 3,683 | 3.1 |  | 19,499 | 1,103 | 5.7 |  | 98,890 | 2,580 | 2.6 |
| 60 to 64 |  | 88,772 | 20 | <0.1 |  | 22,193 | 43 | 0.2 |  | 7,120 | 11 | 0.2 |  | 15,073 | 32 | 0.2 |
| 65 to 69 |  | 43,452 | 1 | <0.1 |  | 10,863 | 0 | <0.1 |  | 4,834 | 0 | <0.1 |  | 6,029 | 0 | <0.1 |
| 70 to 74 |  | 20,604 | 0 | <0.1 |  | 5,151 | 1 | <0.1 |  | 2,773 | 0 | <0.1 |  | 2,378 | 1 | <0.1 |
| 75 to 79 |  | 10,080 | 0 | <0.1 |  | 2,520 | 0 | <0.1 |  | 1,565 | 0 | <0.1 |  | 955 | 0 | <0.1 |
| 80+ |  | 7,864 | 0 | <0.1 |  | 1,966 | 0 | <0.1 |  | 1,483 | 0 | <0.1 |  | 483 | 0 | <0.1 |

*Abbreviations: MHT – Menopausal Hormone therapy*

#### eTable 8. Hospitalisation for endometriosis within 5 years before the index date, by age at index date

|  |  | **Non-MHT initiators** | | |  | **MHT initiators** | | |  | **Starting with unopposed oestrogens** | | |  | **Starting with combined therapy** | | |
| --- | --- | --- | --- | --- | --- | --- | --- | --- | --- | --- | --- | --- | --- | --- | --- | --- |
| **Age group** |  | **N** | **n** | **%** |  | **N** | **n** | **%** |  | **N** | **n** | **%** |  | **N** | **n** | **%** |
| 45 to 49 |  | 523,084 | 5,799 | 1.1 |  | 130,771 | 5,864 | 4.5 |  | 29,538 | 3,679 | 12.5 |  | 101,233 | 2,185 | 2.2 |
| 50 to 54 |  | 1,147,260 | 11,462 | 1.0 |  | 286,815 | 8,546 | 3.0 |  | 50,491 | 5,677 | 11.2 |  | 236,324 | 2,869 | 1.2 |
| 55 to 59 |  | 473,556 | 2,658 | 0.6 |  | 118,389 | 2,171 | 1.8 |  | 19,499 | 1,344 | 6.9 |  | 98,890 | 827 | 0.8 |
| 60 to 64 |  | 88,772 | 199 | 0.2 |  | 22,193 | 172 | 0.8 |  | 7,120 | 115 | 1.6 |  | 15,073 | 57 | 0.4 |
| 65 to 69 |  | 43,452 | 63 | 0.1 |  | 10,863 | 56 | 0.5 |  | 4,834 | 41 | 0.8 |  | 6,029 | 15 | 0.2 |
| 70 to 74 |  | 20,604 | 25 | 0.1 |  | 5,151 | 25 | 0.5 |  | 2,773 | 21 | 0.8 |  | 2,378 | 4 | 0.2 |
| 75 to 79 |  | 10,080 | 8 | 0.1 |  | 2,520 | 10 | 0.4 |  | 1,565 | 8 | 0.5 |  | 955 | 2 | 0.2 |
| 80+ |  | 7,864 | 4 | 0.1 |  | 1,966 | 3 | 0.2 |  | 1,483 | 3 | 0.2 |  | 483 | 0 | <0.1 |

*Abbreviations: MHT – Menopausal Hormone therapy*

#### eTable 9. Infertility treatments within 5 years before the index date, by age at index date

|  |  | **Non-MHT initiators** | | |  | **MHT initiators** | | |  | **Starting with unopposed oestrogens** | | |  | **Starting with combined therapy** | | |
| --- | --- | --- | --- | --- | --- | --- | --- | --- | --- | --- | --- | --- | --- | --- | --- | --- |
| **Age group** |  | **N** | **n** | **%** |  | **N** | **n** | **%** |  | **N** | **n** | **%** |  | **N** | **n** | **%** |
| 45 to 49 |  | 523,084 | 3,342 | 0.6 |  | 130,771 | 6,518 | 5.0 |  | 29,538 | 1,029 | 3.5 |  | 101,233 | 5,489 | 5.4 |
| 50 to 54 |  | 1,147,260 | 1,099 | 0.1 |  | 286,815 | 712 | 0.2 |  | 50,491 | 91 | 0.2 |  | 236,324 | 621 | 0.3 |
| 55 to 59 |  | 473,556 | 67 | <0.1 |  | 118,389 | 42 | <0.1 |  | 19,499 | 3 | <0.1 |  | 98,890 | 39 | <0.1 |
| 60 to 64 |  | 88,772 | 5 | <0.1 |  | 22,193 | 8 | <0.1 |  | 7,120 | 4 | 0.1 |  | 15,073 | 4 | <0.1 |
| 65 to 69 |  | 43452 | 2 | <0.1 |  | 10,863 | 0 | <0.1 |  | 4,834 | 0 | <0.1 |  | 6,029 | 0 | <0.1 |
| 70 to 74 |  | 20604 | 1 | <0.1 |  | 5,151 | 2 | <0.1 |  | 2,773 | 0 | <0.1 |  | 2,378 | 2 | 0.1 |
| 75 to 79 |  | 10080 | 0 | <0.1 |  | 2,520 | 0 | <0.1 |  | 1,565 | 0 | <0.1 |  | 955 | 0 | <0.1 |
| 80+ |  | 7864 | 0 | <0.1 |  | 1,966 | 0 | <0.1 |  | 1,483 | 0 | <0.1 |  | 483 | 0 | <0.1 |

### Treatment patterns among MHT initiators aged 45 years or older

#### eTable 10. Cumulative duration of systemic MHT by age, initiation year, oestrogen administration route, and regimen at initiation

|  | **Number of MHT initiators** | **Median**  **(years)** | **Q1**  **(years)** | **Q3**  **(years)** |
| --- | --- | --- | --- | --- |
| **Overall** | 478,668 | 2.1 | 0.6 | 6.1 |
| **Age at MHT initiation, years** |  |  |  |  |
| 45 – 49 | 130,771 | 1.9 | 0.6 | 6.0 |
| 50 - 54 | 286,815 | 2.6 | 0.8 | 6.8 |
| 55 - 59 | 118,389 | 2.1 | 0.6 | 5.5 |
| 60 - 64 | 22,193 | 1.1 | 0.4 | 3.5 |
| 65 - 69 | 10,863 | 0.9 | 0.4 | 3.0 |
| 70 - 74 | 5,151 | 0.7 | 0.3 | 2.4 |
| 75 - 79 | 2,520 | 0.6 | 0.3 | 1.8 |
| 80+ | 1,966 | 0.5 | 0.3 | 1.3 |
| **Year of MHT initiation** |  |  |  |  |
| 2011 | 60,194 | 2.1 | 0.6 | 5.8 |
| 2012 | 52,877 | 2.0 | 0.6 | 5.7 |
| 2013 | 47,987 | 2.2 | 0.6 | 6.0 |
| 2014 | 48,782 | 1.9 | 0.6 | 5.6 |
| 2015 | 44,708 | 2.0 | 0.6 | 5.7 |
| 2016 | 41,052 | 1.9 | 0.6 | 5.7 |
| 2017 | 38,864 | 2.0 | 0.6 | 5.9 |
| 2018 | 37,498 | 2.1 | 0.6 | 6.2 |
| 2019* | 34,664 | 2.1 | 0.6 | > 6 |
| 2020* | 28,562 | 2.3 | 0.7 | >5 |
| 2021* | 31,263 | 2.3 | 0.7 | >4 |
| 2022* | 31,889 | 2.5 | 0.7 | >3 |
| 2023* | 36,227 | >2 | 0.7 | >2 |
| 2024* | 44,101 | >1 | 0.7 | >1 |
| **Oestrogen administration route at initiation** |  |  |  |  |
| Transdermal | 462,577 | 2.3 | 0.7 | 6.3 |
| Oral | 115,547 | 1.4 | 0.4 | 5.2 |
| Combining oral and transdermal | 544 | 1.3 | 0.5 | 4.7 |
| **MHT regimen at initiation** |  |  |  |  |
| Unopposed oestrogen | 117,303 | 1.4 | 0.5 | 4.8 |
| Oestrogen combined with micronised progesterone or dydrogesterone | 399,855 | 2.6 | 0.7 | 6.6 |
| Oestrogen combined with progestogens other than progesterone or dydrogesterone | 60,961 | 1.4 | 0.5 | 4.5 |

*Abbreviations: MHT – Menopausal Hormone therapy*

**The median or Q3 were not reached for women initiating that year, i.e., they exceed the maximum available follow-up*

#### eTable 11. Prescriber speciality of all systemic oestrogen reimbursements during the follow-up

|  | **Number of reimbursements** | **Proportion** |
| --- | --- | --- |
| **Prescriber speciality** | **N** **= 12,293,418** | **%** |
| General practitioner | 4,952,482 | 40.3 |
| Gynaecologist | 6,835,782 | 55.6 |
| Midwives | 14,788 | 0.1 |
| Other | 490,366 | 4.0 |

### Subgroup Analysis: MHT use among women aged 50-59 years

#### eFigure 6. Prevalence, initiation and discontinuation of systemic MHT among women aged 50-59 years

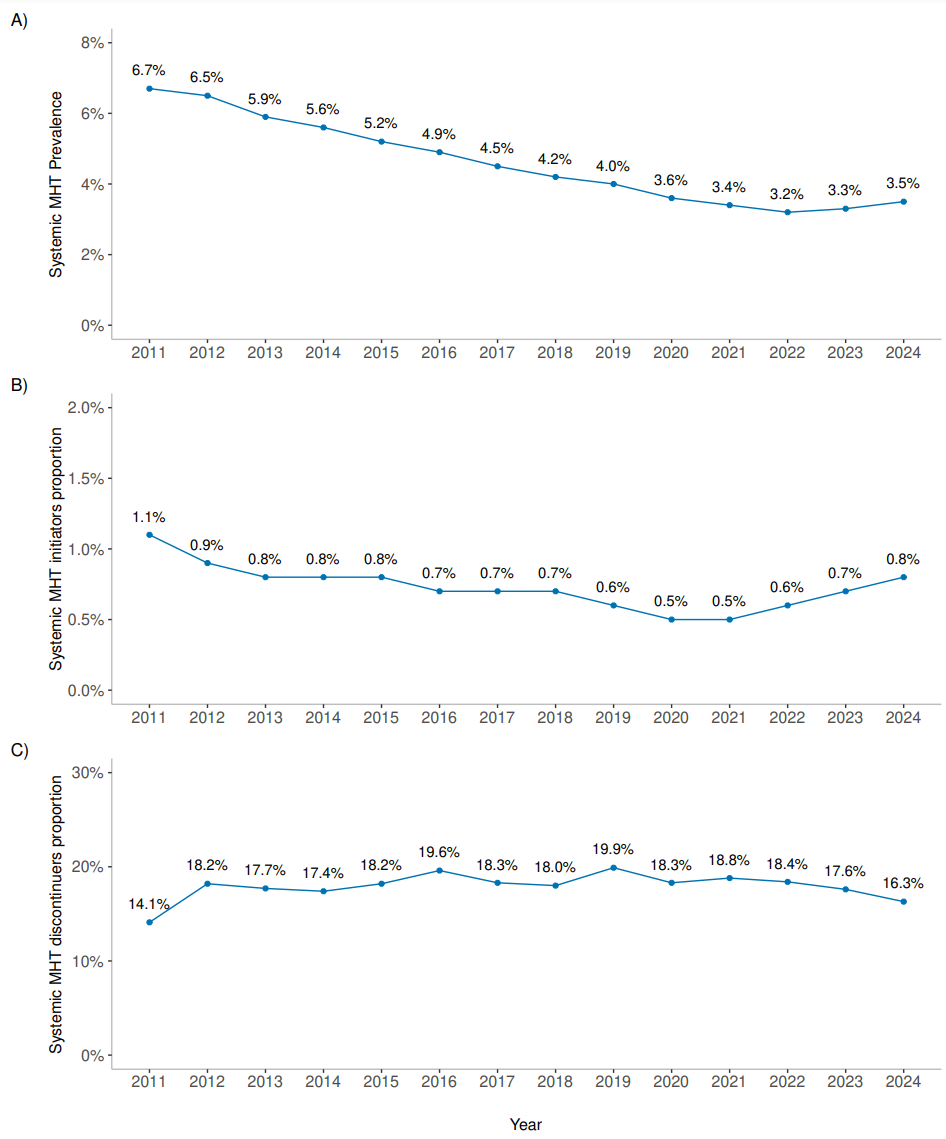

#### eFigure 7. Prevalence of systemic MHT among women aged 50-59 years, by oestrogen administration route and MHT regimen

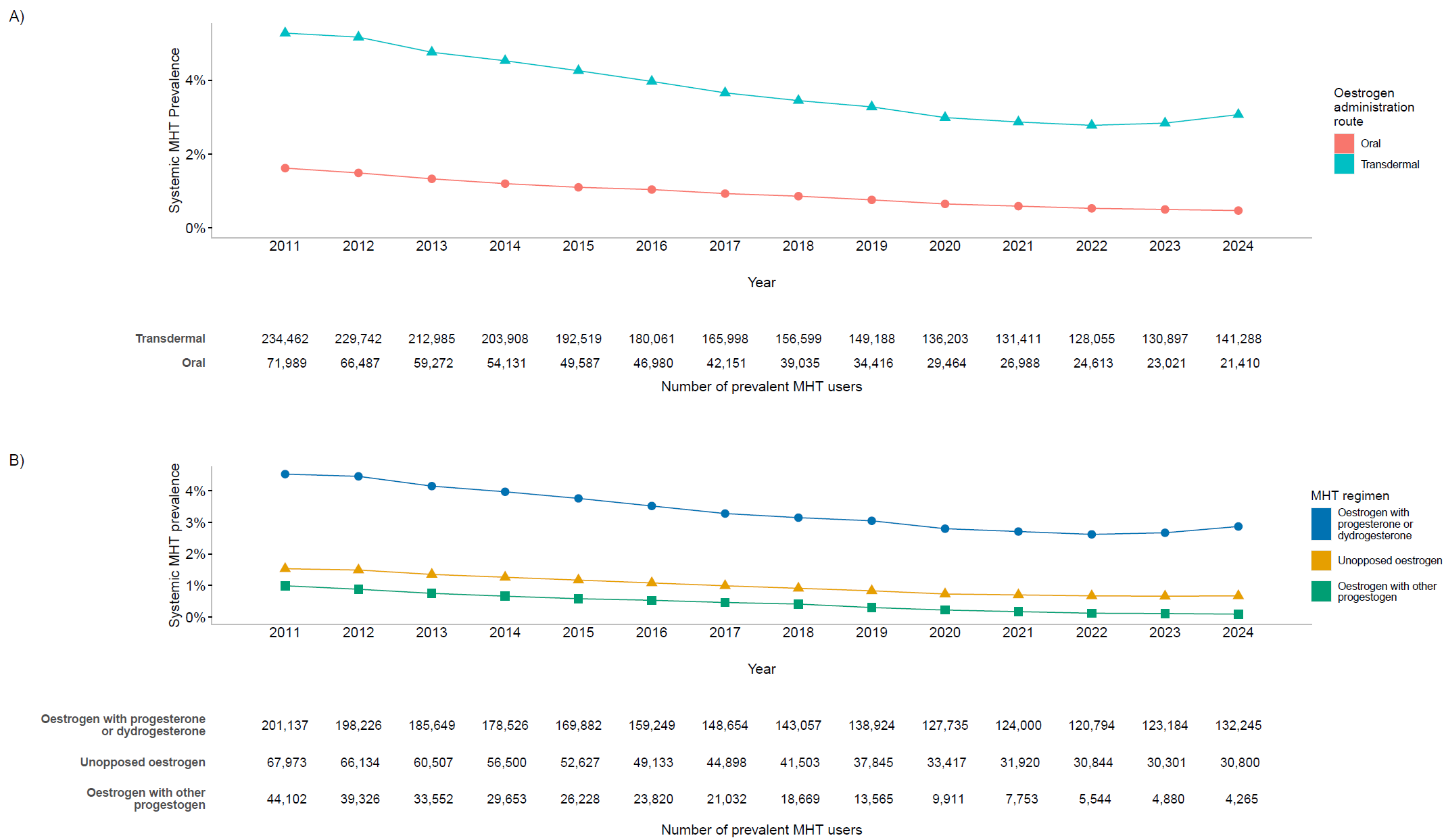

#### eFigure 8. Proportion of systemic MHT initiators among women aged 50-59 years, by oestrogen administration route and MHT regimen

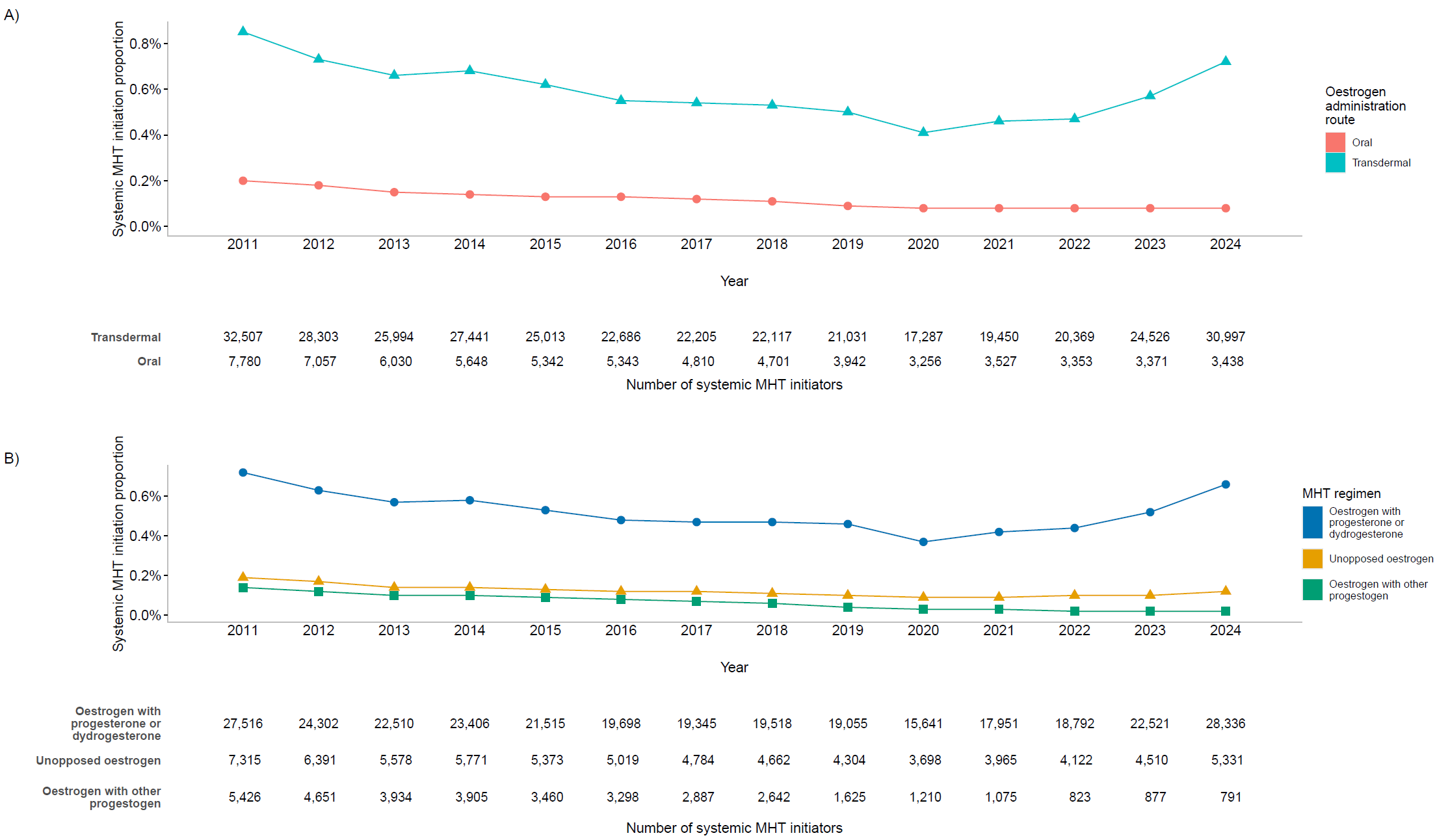

#### eTable 12. Temporal patterns of systemic MHT among initiators aged 50-59 years: treatment duration and treatment episodes

| **Characteristics** |  | **All MHT initiators** | |  | **Starting with unopposed oestrogens** | |  | **Starting with combined therapy** | |
| --- | --- | --- | --- | --- | --- | --- | --- | --- | --- |
| **Cohort, No.** |  | **578,668** | |  | **117,303** | |  | **461,365** | |
|  |  | Median | IQR |  | Median | IQR |  | Median | IQR |
| Total length of the follow-up (years) |  | 8.8 | 4.7 – 12.2 |  | 9.1 | 5.0 – 12.3 |  | 8.8 | 4.7 – 12.2 |
| Time between the first and the last MHT use (years) |  | 3.6 | 1.0 – 8.1 |  | 2.9 | 0.9 – 7.2 |  | 3.8 | 1.1 – 8.2 |
| Cumulative MHT duration (years) |  | 2.4 | 0.7 – 6.4 |  | 1.7 | 0.6 – 5.3 |  | 2.6 | 0.8 – 6.6 |
| Number of MHT episodes |  | 2 | 1 - 4 |  | 2 | 1 - 4 |  | 2 | 1 - 4 |
| Duration of the first MHT episode (years) |  | 1.1 | 0.4 – 3.8 |  | 0.8 | 0.3 – 2.6 |  | 1.2 | 0.4 – 4.1 |
| Duration of the pause between the first two MHT episodes (years)* |  | 0.4 | 0.3 - 0.7 |  | 0.5 | 0.3 - 0.7 |  | 0.4 | 0.3 - 0.7 |

*Abbreviations: MHT – Menopausal Hormone therapy*

** Among 216,344 incident MHT users aged 50-59 for whom at least two exposure episodes were observed*

#### eTable 13. Treatment-related characteristics during follow-up among MHT initiators aged 50-59 years

| **Characteristic** |  | **All MHT initiators** | |  | **Starting with unopposed oestrogens** | |  | **Starting with combined**  **therapy** | |
| --- | --- | --- | --- | --- | --- | --- | --- | --- | --- |
| **Cohort, No.** |  | **405,204** |  |  | **69,990** |  |  | **335,214** |  |
| **Systemic MHT used during follow-up** |  |  |  |  |  |  |  |  |  |
| Treatment regimens |  |  |  |  |  |  |  |  |  |
| Unopposed oestrogens, at least once, n (%) |  | 112,838 | 27.8 |  | 69,990 | 100 |  | 42,848 | 12.8 |
| Combined therapy, at least once, n (%) |  | 351,346 | 86.7 |  | 16,132 | 23.0 |  | 335,214 | 100.0 |
| Unopposed oestrogens, % person-months* |  | 15.4 |  |  | 79.5 |  |  | 3.8 |  |
| Combined therapy, % person-months* |  | 84.6 |  |  | 20.5 |  |  | 96.2 |  |
| Molecule of oestrogen, n (%) |  |  |  |  |  |  |  |  |  |
| Oestradiol only |  | 401,658 | 99.1 |  | 67,409 | 96.3 |  | 334,249 | 99.7 |
| Oestradiol and estriol |  | 1,528 | 0.4 |  | 630 | 0.9 |  | 898 | 0.3 |
| Estriol only |  | 2,018 | 0.5 |  | 1,951 | 2.8 |  | 67 | 0.0 |
| Oestrogen administration routes |  |  |  |  |  |  |  |  |  |
| Transdermal only, n (%) |  | 315,364 | 77.8 |  | 57,750 | 82.5 |  | 257,614 | 76.9 |
| Oral only, n (%) |  | 44,455 | 11.0 |  | 5,813 | 8.3 |  | 38,642 | 11.5 |
| Oral and transdermal, n (%)** |  | 45,385 | 11.2 |  | 6,427 | 9.2 |  | 38,958 | 11.6 |
| Transdermal, % person-months* |  | 84.7 |  |  | 89.2 |  |  | 83.9 |  |
| Oral, % person-months* |  | 15.9 |  |  | 11.5 |  |  | 16.7 |  |
| Oestrogen pharmaceutical forms, n (%) |  |  |  |  |  |  |  |  |  |
| Oral pills only |  | 44,455 | 11.0 |  | 5,813 | 8.3 |  | 38,642 | 11.5 |
| Transdermal patches only |  | 24,496 | 6.0 |  | 6,461 | 9.2 |  | 18,035 | 5.4 |
| Transdermal gels only |  | 256,877 | 63.4 |  | 44,370 | 63.4 |  | 212,507 | 63.4 |
| Oral pills and transdermal patches |  | 4,290 | 1.1 |  | 770 | 1.1 |  | 3,520 | 1.1 |
| Oral pills and transdermal gels |  | 33,661 | 8.3 |  | 4,469 | 6.4 |  | 29,192 | 8.7 |
| Transdermal patches and gels |  | 33,991 | 8.4 |  | 6,919 | 9.9 |  | 27,072 | 8.1 |
| Oral pills and transdermal patches and gels |  | 7,434 | 1.8 |  | 1,188 | 1.7 |  | 6,246 | 1.9 |
| Type of progestogens |  | - |  |  | - |  |  |  |  |
| Micronised progesterone or dydrogesterone, n (%) |  | 302,750 | 74.7 |  | 13,988 | 20.0 |  | 288,762 | 86.1 |
| Progestogens other than progesterone/dydrogesterone, n (%) |  | 21,318 | 5.3 |  | 1,108 | 1.6 |  | 20,210 | 6.0 |
| Both, n (%) |  | 27,278 | 6.7 |  | 1,036 | 1.5 |  | 26,242 | 7.8 |
| None, n (%) |  | 53,858 | 13.3 |  | 53,858 | 77.0 |  | 0 | 0.0 |
| Micronised progesterone or dydrogesterone, % person-months* |  | 79.2 |  |  | 19.2 |  |  | 90.1 |  |
| Progestogens other than progesterone/dydrogesterone, % person-months* |  | 5.3 |  |  | 1.2 |  |  | 6.1 |  |
| Molecule of progestogen (among the 351,346 women using combined therapies at least once) |  |  |  |  |  |  |  |  |  |
| Chlormadinone, n (%) |  | 3,631 | 1.0 |  | 155 | 1.0 |  | 3,476 | 1.0 |
| Cyproterone acetate, n (%) |  | 766 | 0.2 |  | 54 | 0.3 |  | 712 | 0.2 |
| Dienogest, n (%) |  | 197 | 0.1 |  | 4 | 0.0 |  | 193 | 0.1 |
| Dydrogesterone, n (%) |  | 31,943 | 9.1 |  | 1,575 | 9.7 |  | 30,368 | 9.1 |
| Gestodene, n (%) |  | 88 | 0.0 |  | 4 | 0.0 |  | 84 | 0.0 |
| Levonorgestrel, n (%) |  | 1,412 | 0.4 |  | 131 | 0.8 |  | 1,281 | 0.4 |
| Medrogestone, n (%) |  | 691 | 0.2 |  | 47 | 0.3 |  | 644 | 0.2 |
| Medroxyprogesterone acetate, n (%) |  | 665 | 0.2 |  | 45 | 0.3 |  | 620 | 0.2 |
| Nomegestrol, n (%) |  | 3,933 | 1.1 |  | 193 | 1.2 |  | 3,740 | 1.1 |
| Norethisterone acetate, n (%) |  | 3,797 | 1.1 |  | 207 | 1.3 |  | 3,590 | 1.1 |
| Micronised progesterone, n (%) |  | 232,596 | 66.2 |  | 10,868 | 67.0 |  | 221,728 | 66.1 |
| Promegestone, n (%) |  | 1,007 | 0.3 |  | 47 | 0.3 |  | 960 | 0.3 |
| Multiple molecules, n (%) |  | 70,720 | 20.1 |  | 2,902 | 17.9 |  | 67,818 | 20.2 |

*Abbreviations: MHT – Menopausal Hormone therapy*

** Relative to the total number of months of MHT use (rate per 100 months)*

*** Simultaneous or sequential intake*
